# Model-based estimates of age-structured SARS-CoV-2 epidemiology in households

**DOI:** 10.1101/2024.04.18.24306047

**Authors:** Damon J.A. Toth, Theresa R. Sheets, Alexander B. Beams, Sharia M. Ahmed, Nathan Seegert, Jay Love, Lindsay T. Keegan, Matthew H. Samore

## Abstract

Understanding how infectious disease transmission varies from person to person, including associations with age and contact behavior, can help design effective control strategies. Within households, transmission may be highly variable because of differing transmission risks by age, household size, and individual contagiousness. Our aim was to disentangle those factors by fitting mathematical models to SARS-CoV-2 household survey and serologic data. We surveyed members of 3,381 Utah households from January-April 2021 and performed SARS-CoV-2 antibody testing on all available members. We paired these data with a probabilistic model of household importation and transmission composed of a novel combination of transmission variability and age- and size-structured heterogeneity. We calculated maximum likelihood estimates of mean and variability of household transmission probability between household members in different age groups and different household sizes, simultaneously with importation probability and probabilities of false negative and false positive test results. 12.8% of the individual participants showed serologic evidence of prior infection or reported a prior positive test on the survey, and 17.4% of the participating households showed evidence of at least one SARS-CoV-2 importation. Serologically positive individuals in younger age groups were less likely than older adults to have tested positive during their infection according to our survey results. Our model results suggested that adolescents and young adults (ages 13-24) acquired SARS-CoV-2 infection outside the household at a rate substantially higher than younger children and older adults. Our estimate of the household secondary attack rate (HSAR) among adults aged 45 and older exceeded HSARs to and/or from younger age groups. We found lower HSAR in households with more members, independent of age differences. Our findings from age-structured transmission analysis suggest that age groups contact each other at different rates within households, a key insight for understanding community outbreak patterns and mechanisms of differential infection risk.

**Author Summary:** Infectious diseases can spread through human communities in irregular patterns, partly because different demographic groups, such as age groups, experience different transmission risks due to contact or other behavioral or physiological differences. Understanding the factors driving age differences in transmission can help predict patterns of disease spread and suggest efficient public health strategies to mitigate outbreaks. Households are inter-age mixing locations where age differences in transmission can be studied. In early 2021, we collected blood samples from all members of thousands of households in Utah and tested them for SARS-CoV-2 antibodies, from which prior COVID-19 infection can be inferred. We paired these data with mathematical models that quantify probabilities that different combinations of household members end up infected for different assumptions about non-household infection and within-household transmission. Our estimates suggest that adolescents and young adults acquired infection outside the household more frequently than did other age groups. After a household importation occurred, middle-aged and older adults living together transmitted to each other more readily than all other age pairings for a given household size. The age patterns of household transmission we found suggest that within-household contact rate differences play a significant role in driving household transmission epidemiology.

## 1 Introduction

The spread of an infectious disease through a community can occur in complicated patterns, especially when the person-to-person transmission potential of infected individuals is highly variable [1]. Such variability can occur physiologically, through varying duration, severity, and contagiousness of human infections, as well as socially, through varying social settings and contact behaviors that provide transmission opportunities [2]. These variabilities may be partially explained by correlating transmission risk with observable demographic variables such as age and number of contacts in primary social settings such as households [3]. Understanding the mechanisms and associations along which transmission variabilities arise helps improve the efficiency of infection control via targeted pharmaceutical and social interventions [4].

Transmission mechanisms and variabilities could be studied by directly observing the transmissions and transmission opportunities for each infected person, but such data are difficult and costly to collect. Cross-sectional prevalence data, e.g. from testing serological samples for the presence of antibodies among individuals who have had contact with each other, can be used to indirectly estimate transmission rates and patterns. When such data are paired with mechanistically derived mathematical models, insights can be derived about the source of transmission variabilities using appropriate analytic and computational techniques [5, 6]. In this work, our goal was to use such a model-based approach to study transmission variabilities of SARS-CoV-2, with a focus on age-based associations effects, using household seroprevalence data.

SARS-CoV-2 poses high risk of transmission from person to person in indoor public spaces and households [7], where spread of SARS-CoV-2 has been well documented [8]. Studying transmission at the household level is convenient relative to other indoor locations with more transient populations, and household transmission research can provide illuminating insights into risk factors and mechanisms of transmission. Furthermore, households provide an intergenerational mixing ground with contact across age groups, and both household and age structure can be crucially important for accurate characterization of community-wide transmission [3].

Studying differences in susceptibility, transmissibility, and contact by age has been an important area of research with implications for public health policy for many infectious diseases. During the COVID-19 pandemic, younger individuals have been less likely to experience severe disease but are a potential source of SARS-CoV-2 contagion [9]. Infected children with mild or no symptoms may nevertheless transmit to other more vulnerable contacts, including to highly vulnerable aged adults in multi-generational households [10]. Some SARS-CoV-2 and influenza studies have found that households with more members have lower transmission rates [11-13], and it is unclear the extent to which that observation might be attributed to different age configurations of larger households or other factors. Understanding differences in transmission among and between different age groups, and the mechanisms by which those differences might arise, can shed light on the benefits of age-group targeting of transmission-interruption measures, including vaccination [14].

Serological surveys, in which blood samples from individuals in a population are tested for the presence of antibodies, can be used to infer which individuals had experienced prior infection, including those who had not been diagnosed [15]. When performed over entire households, serological surveys offer one way to assess household transmission: the number of household members testing positive provides information to infer the distribution of the total number of household members infected during household outbreaks that ran their course prior to data collection. Quantitative model-based techniques [16] can be used to extract information from these datasets on the transmissibility of the pathogen under conditions of sustained close contact. When such datasets include metadata on household member demographics, they can help answer questions about the relative susceptibility to or transmissibility of infection across demographics such as age [17].

In this work, we developed a novel mathematical model of household transmission that incorporates transmission variability [18] and heterogeneity over age and household size. A central goal achieved by our model-based findings was to estimate quantitative age differences in household COVID-19 epidemiology using detailed serological data from Utah households and elucidate the extent to which these estimates reveal potential mechanisms of within-household transmission differences by age.

## 2 Methods

### 2.1 Data collection from Utah households

Our data was collected via the Utah Health & Economic Recovery Outreach (HERO) Project [19], which recruited participants from households in the state of Utah during several distinct phases in 2020 [20] and 2021 [21]. Members of participating households completed surveys and were subsequently invited to a mobile testing site to contribute samples for testing. Participants contributing the specific subset of the Utah HERO Project data analyzed and modeled for this manuscript were recruited during two different phases of the Utah HERO Project in 2021: “school district testing” and “hotspot testing” (see Supporting Information for recruitment details). In contacted households, all household members aged 12 years or older were asked to complete an individual survey, adults were asked to complete the individual survey on behalf of children <12 years of age, and the household was asked to complete an overall household survey. Each household member >6 months of age was asked to provide a blood sample at a mobile testing site.

The University of Utah Institutional Review Board reviewed the surveillance project that produced the data analyzed in this manuscript and determined it as non-research public health surveillance, waived the requirement for documented consent, and determined that use of these data for analysis to understand the dynamics of SARS-CoV-2 transmission was exempt from further review (IRB_00132598). Individuals were informed of the project procedures and that participation was voluntary. Participants provided their agreement to participate and were given the chance to opt out of having their data used for future research. The data were analyzed anonymously for this manuscript.

The following data from the surveys were retained for our analysis. From the household survey, we used the household size (number of people living in the household). From the individual surveys, we used the date of birth, whether the individual had previously tested positive for COVID-19, the date of a previous positive test, the number of prior COVID-19 vaccine doses received, and the date of any prior vaccine doses. We calculated the age of surveyed participants on the day of serological testing, using the date of birth from the survey. Prior test result data from the survey were classified as prior positive reported (P) or no prior positive reported (N).

We excluded households from our analysis if the number of people living in the house (household size) or the age of any household member were missing. We also excluded households for which the reported size was inconsistent with the individual-level data (i.e., the reported household size was less than the number of individuals contributing data for that household).

Whole blood was collected from participants at mobile testing facilities, via finger prick for participants aged between 6 months and 12 years and via venous puncture for participants aged 12 years or older. Serum specimens from blood collected via venous puncture were analyzed using all three of the following tests: Abbott SARS-CoV-2 IgG assay performed on an Abbott Architect i2000 instrument (Abbott Laboratories), Euroimmun Anti-SARS-CoV-2 ELISA (IgG) assay, and Siemens SARS-CoV-2 IgG (Cov2G) assay. Only the Euroimmun test was used to analyze specimens from finger prick samples.

Methodology and criteria for a positive antibody result were defined according to the manufacturers’ instructions [22-24] and as in prior studies [25, 26]. Antibody test results for each test were classified according to negative (N), positive (P), or not performed / indeterminate (X). Because the Euroimmun and Siemens assays detect anti-SARS-CoV-2 IgG for the S1 domain of viral spike protein (anti-S1), which can be produced by vaccination, participants who reported having received at least one dose of COVID-19 vaccine prior to data collection and tested positive on either assay were automatically labeled as indeterminate (X) for purposes of informing prior infection. The Abbott assay detects anti-SARS-CoV-2 IgG for the nucleocapsid protein (anti-N), which we assumed was not impacted by prior vaccination and that vaccinated patients testing Abbott-positive had tested positive for prior infection [27]. We defined an antibody test result as being “informative” for potential prior infection status when the result was negative or positive for the Abbott test, negative or positive for Euroimmun or Siemens for participants reporting no prior vaccination, or negative for Euroimmun or Siemens for participants reporting any prior vaccination.

Each member of a household retained for analysis was labeled according to their combination of five data points: reported prior positive test (N or P); Abbott test result (N, P, or X); Euroimmun test result (N, P, or X); Siemens test result (N, P, or X). For example, a participant labeled “NPXN” reported no prior positive test, tested positive by Abbott, had an indeterminate Euorimmun test, and tested negative by Siemens. Further labels were added indicating age group and household size, as described in later sections. Then we created a table tallying the number of participants with each label in each household: a column for each unique label that at least one participant received and a row for each household. Each table element is the number of participants in each household with each label. We then tallied the number of unique rows, *C*, and labeled the sets of numbers in each unique row as vectors **y**_1_, …, **y**_*C*_. Finally, the entire dataset is represented by the vector **y** = (**y**_1_, …, **y**_*C*_, *f*_1_, … *f*_*C*_), where frequency *f*_*i*_ is the number of households described by the corresponding **y**_*i*_.

The following sections describe the mathematical model designed to produce a likelihood function ℒ(**y**) to quantify the probability of observing the dataset **y** under a given set of epidemiological assumptions. The de-identified data and codes, written in R version 4.0.3, to produce **y**, calculate and optimize ℒ(**y**), and produce the results in this manuscript, are publicly available at https://github.com/damontoth/householdTransmission2021.

### 2.2 Household transmission model

First, we define the probability that a single infected person transmits infection to different numbers of people living in the same household. We separate a household’s population into age groups that can differ in both susceptibility to acquisition from infected housemates and in transmissibility to susceptible housemates from each group. For simplicity, we limited the number of transmission age groups to three, named U, V, and W. We use the notation ***F***_**y**;**z**_ to denote the probability that a single infected person with **z** = (z_u_, z_v_, z_w_) susceptible housemates in each group directly transmits infection to **y** = (*y*_u_, *y*_v_, *y*_w_) of them, respectively.

To derive our formula for ***F***, we assume that the probability of transmission to a u-group person is a random variable taking the form 1 *− e*^*−x*^, where *x* is a gamma-distributed random variable with shape *k* and rate *r*. The gamma distribution models the mean and variability of infected individuals’ transmissibility to housemates, where the *k* parameter is equivalent to the dispersion parameter *k* often used to model transmission variability and superspreading. The value of *x* can be considered the product of the total dose of exposure experienced by an infected person’s susceptible housemate and the per-dose probability of infection, as in the exponential dose-response model. Then, the probabilities of transmission to a person in group V and W is 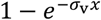 and 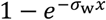, where the coefficients *σ*_v_ and *σ*_w_ alter the probability of infection relative to a person in group U, from a given infected housemate.

From the above assumptions, the following formula for ***F*** applies (see Supporting Information for derivation):

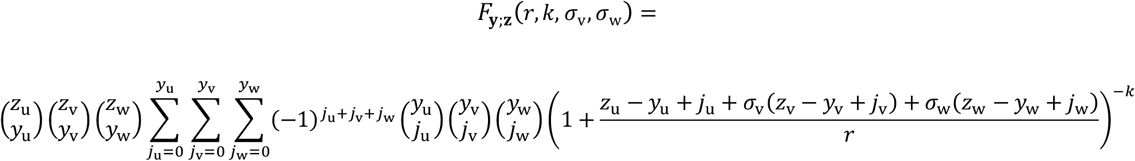

Because we also model differential transmissibility from each group, we use three sets of (*r, k, σ*_v_, *σ*_w_) parameters governing transmission: (*r*_u_, *k*_u_, *σ*_uv_, *σ*_uw_), (*r*_v_, *k*_v_, *σ*_vv_, *σ*_vw_), and (*r*_w_, *k*_w_, *σ*_wv_, *σ*_ww_), which are the parameters used when a person from group U, V, and W is the potential transmitter, respectively. Using those parameters, we can also define the directional household secondary attack rates (HSARs) *p*_*ij*_, the mean transmission probability from an infected person from group *i* to a susceptible person from group *j* in the same household:

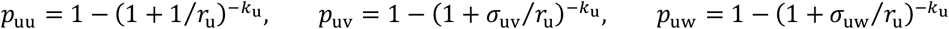

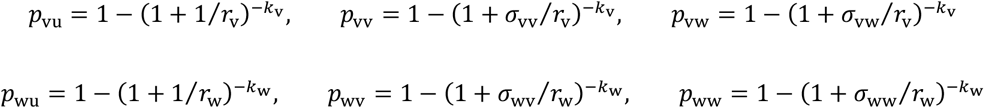

For clarity, we report our estimates for the set of transmission parameters using the above nine directional attack rates, plus the dispersion parameters *k*_*i*_. The values of *r*_*i*_ and *σ*_*ij*_ can then be calculated by inverting the above equations.

With the above function ***F*** defined for transmissions from one infectious individual in a household, we now derive formulae for multiple potential transmitters. First, let *H*_**x**;**y**;**z**_ be the probability that, in a household with **x** = (*x*_u_, *x*_v_, *x*_w_) infected members from each group, those infected persons transmit infection directly to a total of **y** = (*y*_u_, *y*_v_, *y*_w_) of **z** = (z_u_, z_v_, z_w_) susceptible housemates from each group. First, we define the trivial case where **x** = (0,0,0): no infected household members to transmit:

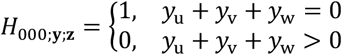

Next, we have already defined the cases where *x*_u_ + *x*_v_ + *x*_w_ = 1 (exactly one infected household member):

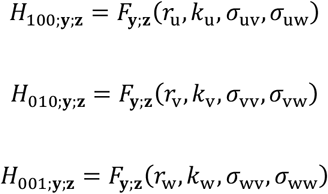

Then we iteratively calculate *H*_**x**;**y**;**z**_ for each (*x*_u_, *x*_v_, *x*_w_) in increasing sequence of *x*_u_ + *x*_v_ + *x*_w_ = 2, 3, …:

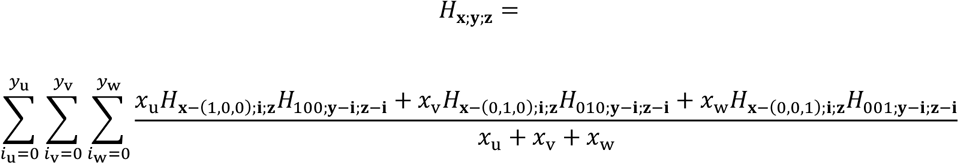

With *H* in hand, we then calculate *T*_**x**;**y**;**z**_: the probability that **x** already infected (u-group, v-group, w-group) household members lead to a *total* of **y** transmissions to **z** susceptible household members from each respective group. In other words, *T*_**x**;**y**;**z**_ is the probability that the vector of final household outbreak sizes of each group is **x** + **y** given that **x** were initially infected and **z** were initially susceptible. The probability accounts for transmissions directly from initially infected members and in any subsequent transmission generations.

We calculate values of *T* iteratively, starting with trivial case of no initial infections in the household:

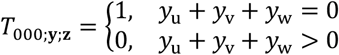

Then, starting with *y*_u_ + *y*_v_ + *y*_w_ = 0 and then for increasing *y*_u_ + *y*_v_ + *y*_w_ in sequence, we iteratively calculate:

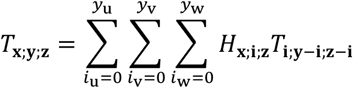

### 2.3 Total household infection size model

The probability of infections within a household also depends on the likelihood of members acquiring infection from non-household members (called “community” acquisitions). We calculated those probabilities for each household member based on characteristics that could extend beyond membership in the U, V, and W transmission groups described in the last section.

Let **n** = (*n*_u_, *n*_v_, *n*_w_) be the number of household members in each of the three transmission groups. Let *l*_1_, …, *l*_*n*_ be the infection indicators for each household member, where the *l*_*i*_ values equal 1 if infected and 0 if uninfected. Let *u*_1_, …, *u*_*n*_ be indicators for whether each household member belongs to transmission group U (1 if yes, 0 if no), and similarly *v*_1_, …, *v*_*n*_ and *w*_1_, …, *w*_*n*_ are indicators for membership in groups V and W. Then we define 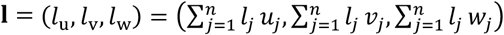.

Let *p*_1_, …, *p*_*n*_ be the community acquisition probability of each household member. Now we define the likelihood 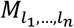 for household members indicated by the *l* values ending up infected by either community acquisition or household transmission.

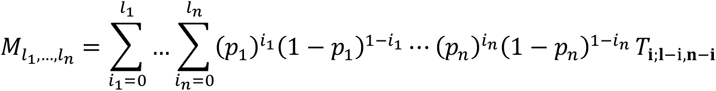

The values 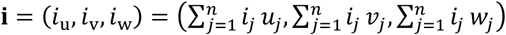. The *M* values depend on both the household transmission parameters defined in the prior section and the community acquisition probabilities.

### 2.4 Data accuracy model

Applying the *M* formula from the prior section directly to our data would be problematic because the true infection history of each household member is not known with certainty. Our data include up to 4 sources of COVID-19 test information by which prior infection status of individual household members can be probabilistically inferred: results from up to 3 different antibody tests (Abbot, Siemens, and Euroimmun) and results from surveys in which participants could report results of a prior test.

Antibody test results are subject to imperfect sensitivity and specificity due to false negatives and false positives. To account for these, we added 6 additional parameters to our likelihood model: *ϕ*_*AA*_, *ϕ*_*AS*_, and *ϕ*_*AE*_ are the antibody test sensitivities for Abbot, Siemens, and Euroimmun, respectively, and π_*AA*_, π_*AS*_, and π_*AE*_ are the specificities. Prior test results for SARS-CoV-2 reported on the survey also do not perfectly identify those with prior infections. Among people who had a prior infection, a portion of them never tested positive because they did not receive a test while their infection was identifiable or because they falsely tested negative. Because the degree of this under-ascertainment has been known to vary widely by age, we modeled it using a separate probability for each household member: (*ϕ*_*S*1_, …, *ϕ*_*Sn*_) are the probabilities that each of the *n* surveyed persons in a household reported receiving a positive test for the virus had they been infected. The parameter π_*S*_ is the probability that any surveyed person with no prior infection did not report receiving a positive test.

For each person *i* = 1, …, *n* in a household of size *n*, we considered the overall combination of that person’s test results to calculate *ϕ*_*i*_ and π_*i*_, the probability of that test result combination occurring if the person had been infected and never infected, respectively. The following provide illustrated examples.

Example 1: result combination “NNNN”: no prior positive reported on survey, and negative by all 3 antibody tests:

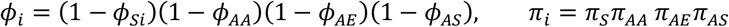

Example 2: result combination “NNPN”: no prior positive reported on survey, negative by Abbot, positive by Euroimmun, and negative by Siemens:

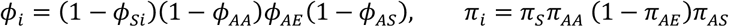

Example 3: result combination “PNXN”: prior positive reported on survey, negative by Abbot, no or indeterminate Euroimmun result, and negative by Siemens:

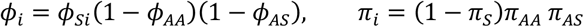

With the *ϕ*_*i*_ and π_*i*_ values for each household member in hand, the following formula specifies the overall likelihood of observing the entire household’s set of test results together:

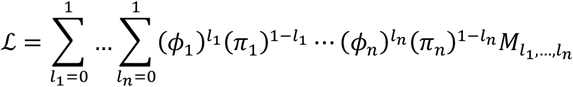

### 2.5 Age- and size-based parameter values

The formulas in the previous sections included three different components that we assumed could depend on the characteristics of household member *i*. First, the “transmission group” specified by (*u*_*i*_, *v*_*i*_, *w*_*i*_); second, the community acquisition probability *p*_*i*_; and third, the prior infection ascertainment probability *ϕ*_*Si*_. We modeled each of these dependencies based on the individual’s age at the time of data collection. Specifically, we categorized ages using combinations of age ranges for terms defined by Medical Subject Headings (MeSH) [28] (Table 1). Because we had limited data from the youngest individual age groups, we combined the newborn, infant, and preschool child groups into one category (aged 0 to 5 years). Other MeSH-defined age categories and sub-categories were retained separately, for a total of 8 categories.

**Table 1.**
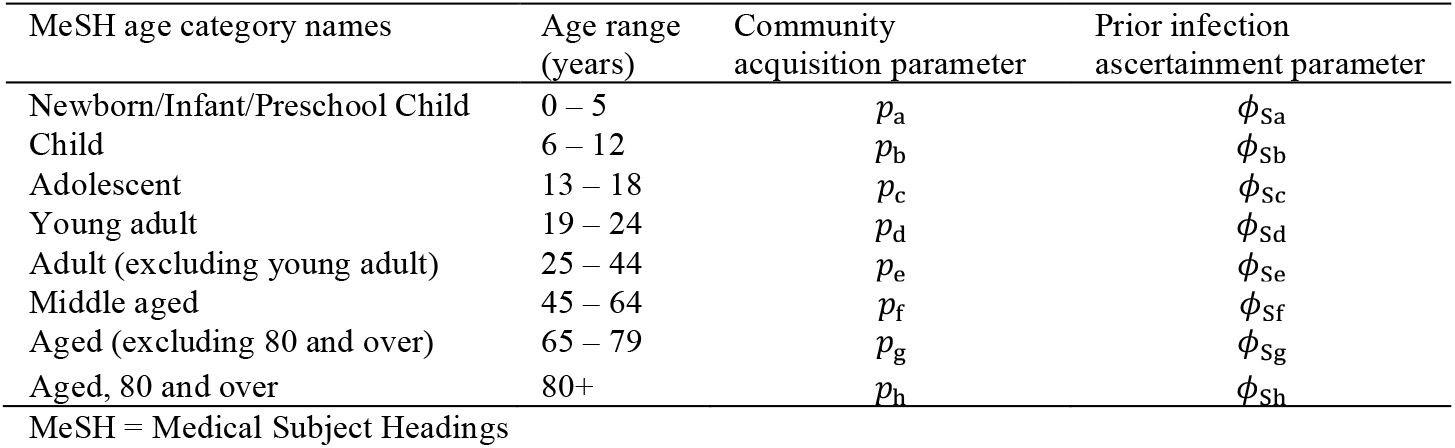
Age categories and age-specific model parameters.

| MeSH age category names | Age range (years) | Community acquisition parameter | Prior infection ascertainment parameter |
| --- | --- | --- | --- |
| Newborn/Infant/Preschool Child | 0 – 5 | $p_a$ | $\phi_{Sa}$ |
| Child | 6 – 12 | $p_b$ | $\phi_{Sb}$ |
| Adolescent | 13 – 18 | $p_c$ | $\phi_{Sc}$ |
| Young adult | 19 – 24 | $p_d$ | $\phi_{Sd}$ |
| Adult (excluding young adult) | 25 – 44 | $p_e$ | $\phi_{Se}$ |
| Middle aged | 45 – 64 | $p_f$ | $\phi_{Sf}$ |
| Aged (excluding 80 and over) | 65 – 79 | $p_g$ | $\phi_{Sg}$ |
| Aged, 80 and over | 80+ | $p_h$ | $\phi_{Sh}$ |

MeSH = Medical Subject Headings

The community acquisition parameter for each individual *p*_*i*_ in a household was set to one of the 8 values *p*_a_ through *p*_h_ according to age group, and similarly each individual’s prior infection ascertainment parameter *ϕ*_S*i*_ was set to a value among *ϕ*_Sa_ through *ϕ*_Sh_. For the transmission groupings, because we limited our model to three groups, we tested different ways to combine our 8 MeSH age groups from Table 1 into three groups of adjacent MeSH age groups. We designated transmission group “U” as the oldest age group, “V” as the middle age group, and “W” as the youngest.

We also allowed within-household transmission to vary by total household size (number of household members) using two additional parameters: *σ*_34_ and *σ*_5+_ scale the probability of acquiring infection from an infected housemate of a person living in a size 3-4 or size 5-or-more household, respectively, relative to a person living in a size-2 household. These parameters multiply each of the *σ*_xy_ parameters that model age differences, so that each age-to-age transmission probability is assumed to scale by household size in the same way. Thus, we calculate three sets of *T* values quantifying household transmission chain probabilities, for a given set of epidemiological parameters:

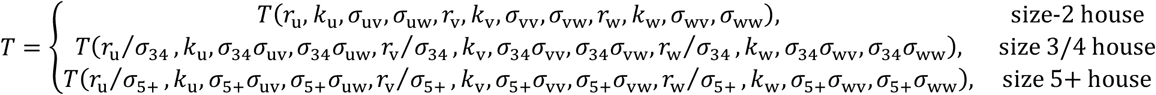

In our full model we sought to use our data to simultaneously estimate the 22 epidemiological parameters

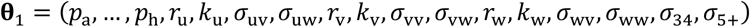

and the 15 data accuracy parameters

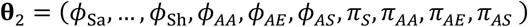

using maximum likelihood estimation (MLE).

The log likelihood of the dataset **y** described in Section 2.1 with variable set **θ** = (**θ**_1_, **θ**_2_) is

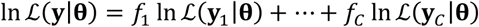

### 2.6 Likelihood optimization and uncertainty

We maximized the log likelihood over the unknown parameters **θ** using the observations for each household, to produce the MLE 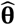. The log likelihood maximization was performed using an iterative scheme in R, in which R’s native “optim” function was applied alternately to the set of epidemiological parameters **θ**_1_ and then to the set of data accuracy parameters **θ**_2_, holding the other set constant at the previous solution and continuing back and forth until the estimates converged.

We first tested models with simpler assumptions for the dispersion parameters *k*_u_, *k*_v_, and *k*_w_. The value *k* = 1 is a special case representing exponentially-distributed (special case of gamma-distributed) transmissibility by an infected person. The special case *k* → ∞ represents no variance in transmissibility. We tested models assuming *k*_u_ = *k*_v_ = *k*_w_ = 1 and *k*_u_ = *k*_v_ = *k*_w_ = ∞ against one that optimized those values along with other parameters; if one of the simpler models is favored, we could move forward with other model comparisons described next using less computationally costly formulae.

We also tested different age-group choices for the household transmission groups U, V, and W described above, and we selected the grouping that produced the MLE with the highest likelihood. We refer to this result as the MLE of the “full model” to which alternate, simpler models were compared, to test the extent to which the full model MLE might suffer from overfitting.

For the full model, we derived approximate confidence interval boundaries for an individual parameter *θ*_*i*_ using the likelihood ratio test, using the statistic 2 log 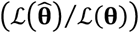, where **θ** consists of *θ*_*i*_ freely varying and the other elements of **θ** held at their optimal value. We defined a 95% confidence interval boundary where *θ*_*i*_ produces a value for this statistic equal to the 95^th^ percentile of the chi-squared distribution with 1 degree of freedom.

### 2.7 Alternate model comparisons

We compared the MLE of the full model against simpler models with fewer free parameters. To calculate P-values at which a simpler model could be rejected in favor of one with more free parameters and of which the simpler model is a special case, we used the chi-squared distribution with degrees of freedom equal to difference in the number of free parameters between the two models. We quantified and compared the parsimony of different models using the Akaike information criterion, which aims to balance goodness of fit with model simplicity.

For the set of average HSARs, which consists of 9 free age-group-to-age-group values in the full model, we tested alternate models that use fewer parameters via simplifying assumptions. We tested an “equal contact” model, which assumes that the extent to which household members contact each other is independent of age, but that susceptibility and/or transmissibility per contact can differ by age. The equal contact model corresponds to the assumption that *σ*_uv_ = *σ*_vv_ = *σ*_wv_ and *σ*_uw_ = *σ*_vw_ = *σ*_ww_. Thus, in this model the 9 HSARs are governed by 5 free parameters: 2 relative susceptibility (*σ*) parameters and 3 average transmissibility parameters.

Next, we tested a “reciprocal transmission” model, which assumes that the average household transmission probability between an infected and susceptible person from two different age groups is the same regardless of which person is infected, i.e., *p*_vu_ = *p*_uv_, *p*_wu_ = *p*_uw_, and *p*_wv_ = *p*_vw_. With those 3 constraints, the reciprocal transmission model is governed by 6 free parameters to determine the 9 HSARs.

To seek a maximally parsimonious model, we tested an additional series of models applying a variety of simplifying assumptions, as defined by optimal AIC. These included assumptions of age-independence of transmission to and/or from each of the individual transmission age groups. For example, assuming *p*_uu_ = *p*_uv_ = *p*_uw_ enforces transmission from age group U to be equally likely to a housemate of any age. Likewise, assuming *p*_uu_ = *p*_vu_ = *p*_wu_ enforces acquisition of infection by age group U to be equally likely from an infected housemate of any age, and assuming *p*_uu_ = *p*_uv_ = *p*_uw_ = *p*_vu_ = *p*_wu_ enforces an equal HSAR between age group U and all ages in both directions. We repeated the MLE procedure for each of these models and each age group, as well as combinations of them for multiple age groups in cases where multiple sets of assumptions produced favorable fits by the AIC. We also tested models that collapse the different age-group-based community acquisition parameters *p*_a_ through *p*_h_ and prior infection ascertainment parameters *ϕ*_Sa_ through *ϕ*_Sh_ into fewer age groupings, as well as models that assume no effect of household size on transmission rates. We combined any of the above simplifications that were favored by the AIC criteria to arrive at our maximally parsimonious model.

Finally, we analyzed implications of our results for the most parsimonious model. We explored potential mechanisms of household transmission difference by age that are consistent with estimated age differences in HSAR, including age-specific differences in household contact rates and per-contact transmissibility or susceptibility. We also calculated the overall age-specific infection risk for each group implied by the model results and the age configurations of households in the data to provide a summary of the estimated rate of infection from household and non-household sources for each age group.

## 3 Results

### 3.1 Data summary

Of the 96,547 total households contacted across both the school district and hotspot testing phases, we compiled serological test data and associated survey data from 3,381 (3.5%) of the households and 7,707 individuals (Table A in Supporting Information).

Of the 3,381 total households, we retained 2,514 (74.4%) for our analysis. The 867 excluded households were removed because of unknown household size (n=55), a reported household size inconsistent with individual data (n=57), or incomplete age data obtained for the household members (n=755). In the 2,514 retained households, there were 5,855 (2.33 per household) household members.

Among the 5,855 retained participants, 1,337 (22.8%) reported having received at least one dose of a COVID-19 vaccine prior to serological data collection, and 727 (12.4%) reported having received two vaccine doses. For those reporting a plausible date for their first vaccine dose (n = 1274), the reported date occurred a median of 32.5 days prior to serological sample collection (range 0–112 days). For those reporting plausible dates for two vaccine doses (n = 654), the reported date of the second dose occurred a median of 29 days prior to data collection (range 0–91 days). Participants reporting prior vaccination tested antibody positive by Euroimmun and Siemens tests at highly elevated rates compared to those not reporting prior vaccination (Table B in Supporting Information), consistent with the presence of vaccine-induced antibodies; these positive results were labeled non-informative (‘X’) when calculating prior SARS-CoV-2 infection likelihood in our model.

5,626 (96.1%) of the retained participants provided a blood sample that produced at least one informative antibody test result, and 2,848 (48.6%) of the retained participants received informative results for all three antibody tests. Of those, 2,412 (84.7%) were all three negative, 230 (8.1%) were all three positive, and 206 (7.2%) were a mixture of positive and negative results. The most common conflicting result combinations among the latter group were Abbott-negative conflicting with Euroimmun-positive and Siemens-positive results (n=99) and Euroimmun-positive conflicting with Abbott-negative and Siemens-negative results (n=63). We tallied other result combinations, including indeterminate (non-informative) results marked by ‘X’ as well as prior-positive test reporting for participants in each antibody test result group (Table C in Supporting Information).

In all, 748 (12.8%) of the 5,855 participants were positive on at least one informative antibody test or reported prior test. These 748 participants lived in 438 of the 2514 households, thus 17.4% of the participating households had at least one informative positive result. Among those 438 households with at least 1 informative positive participant,

246 had exactly 1 and 192 had more than 1, ranging from 2 to 8. A crude HSAR estimate, i.e., assuming exactly 1 importation in each of the 438 households and calculating the fraction of housemates of an importer who were also informative positive, was 33.9%. Our likelihood model relaxed those assumptions in several ways, including accounting for the probability of multiple household importations, false negative and false positive tests, and other features, as detailed in the following sections.

### 3.2 Maximum likelihood estimates

#### 3.2.1 Preliminary model comparisons: transmission dispersion and age groupings

We compared models that produced estimates of each individual transmission dispersion parameter (*k*_u_, *k*_v_, *k*_w_) to simpler models of transmission variability. First, the simpler model with exponentially distributed transmissibility (*k*_u_ = *k*_v_ = *k*_w_ = 1) produced similar maximal likelihood to the full model and a superior AIC, while the model with no variability in transmissibility (*k*_u_ = *k*_v_ = *k*_w_ = ∞) produced inferior maximal likelihood (Table D in Supporting Information). For simplicity we fixed the most parsimonious *k*_u_ = *k*_v_ = *k*_w_ = 1 assumption for all following models.

We also ran our MLE procedure for each possible configuration of the (U, V, W) transmission groups (Table E in Supporting Information). The best configuration producing the maximal log-likelihood occurred when the oldest group, reference group U, included middle aged and aged individuals (age 45+), the middle group V included adults excluding young adults (age 25-44), and the youngest group W included children, adolescents, and young adults (age 0-24). We used those three age ranges for the three transmission age groups in all following models.

#### 3.2.2 Full model results

The model with the assumptions defined in the prior sub-section and no other simplifying assumptions is called our “full model” and includes 34 free parameters. We derived the MLE and confidence intervals for all 34 parameters (Table 2).

**Table 2:** Full model MLE results with confidence intervals.

| Parameter | MLE (95% CI) | 394 |
| --- | --- | --- |
| Mean community acquisition probability |  |  |
| – infant/pre-school 0-5 ($p_a$) | 3.2% (0.6% – 6.8%) | |
| – child 6-12 ($p_b$) | 2.0% (0.6% – 4.1%) | |
| – adolescent 13-18 ($p_c$) | 15.1% (11.4% – 19.4%) | |
| – young adult 19-24 ($p_d$) | 12.6% (8.8% – 17.2%) | |
| – adult 25-44 ($p_e$) | 6.7% (5.5% – 8.0%) | |
| – middle aged 45-64 ($p_f$) | 6.7% (5.3% – 8.4%) | |
| – aged 65-79 ($p_g$) | 5.2% (3.7% – 7.1%) | |
| – aged, 80 and over ($p_h$) | 0.0% (0.0% – 3.4%) | |
| HSAR to age 45+ adults in 2-person households |  |  |
| – From middle aged and older adults aged 45+ ($p_{uu2}$) | 68% (56% – 77%) | |
| – From adults aged 25-44 ($p_{vu2}$) | 45% (11% – 66%) | |
| – From children, adolescents, and young adults aged 0-24 ($p_{wu2}$) | 42% (23% – 57%) | |
| HSAR to age 25-44 adults in 2-person households |  |  |
| – From middle aged and older adults aged 45+ ($p_{uv2}$) | 52% (22% – 73%) | |
| – From adults aged 25-44 ($p_{vv2}$) | 44% (29% – 58%) | |
| – From children, adolescents, and young adults aged 0-24 ($p_{wv2}$) | 63% (50% – 74%) | |
| HSAR to age 0-24 individuals in 2-person households |  |  |
| – From middle aged and older adults aged 45+ ($p_{uw2}$) | 34% (11% – 53%) | |
| – From adults aged 25-44 ($p_{vw2}$) | 47% (31% – 60%) | |
| – From children, adolescents, and young adults aged 0-24 ($p_{ww2}$) | 45% (33% – 56%) | |
| Altered transmission coefficient to members of larger households |  |  |
| – with 3 or 4 members ($\sigma_{34}$) | 0.43 (0.33 – 0.57) | |
| – with 5 or more members ($\sigma_{5+}$) | 0.22 (0.16 – 0.29) | |
| Probability that prior infected person reported receiving a prior positive test |  |  |
| – infant / pre-school, 0-5 ($\phi_{sa}$) | 16% (6% – 32%) | |
| – child, 6-12 ($\phi_{sb}$) | 36% (23% – 49%) | |
| – adolescent, 13-18 ($\phi_{sc}$) | 53% (42% – 63%) | |
| – young adult 19-24 ($\phi_{sd}$) | 62% (47% – 75%) | |
| – adult 25-44 ($\phi_{se}$) | 59% (52% – 65%) | |
| – middle aged 45-64 ($\phi_{sf}$) | 73% (65% – 80%) | |
| – aged 65-79 ($\phi_{sg}$) | 73% (61% – 83%) | |
| – aged, 80 and over ($\phi_{sh}$) | 100% (38% – 100%) | |
| Probability that prior infected person (any age) tested positive for antibodies: |  |  |
| – Abbot ($\phi_{AA}$) | 72% (68% – 76%) | |
| – Euroimmun ($\phi_{AE}$) | 99.2% (97.5% – 99.98%) | |
| – Siemens ($\phi_{AS}$) | 87% (83% – 90%) | |
| Probability that uninfected person did not report receiving a prior positive test |  |  |
| – any age ($\pi_s$) | 99.2% (98.9% – 99.4%) | |
| Probability that uninfected person (any age) tested negative for antibodies |  |  |
| – Abbot ($\pi_{AA}$) | 99.7% (99.4% – 99.8%) | |
| – Euroimmun ($\pi_{AE}$) | 98.1% (97.6% – 98.6%) | |
| – Siemens ($\pi_{AS}$) | 99.85% (99.65% – 99.95%) | |

For community acquisition probability, the highest estimates occurred for adolescents: *p*_*c*_ = 15.1% (11.4% – 19.4%) and young adults: *p*_*d*_ = 12.6% (8.8% – 17.2%). The confidence intervals for those estimates did not overlap with those of any other age group, younger or older. The community acquisition probability for children aged 6–12: *p*_*b*_ = 2.0% (0.1% – 4.1%) was substantially lower than the adjacent age group of adolescents, and *p*_*b*_ was also lower than the result for adults aged 25-44: *p*_*e*_ = 6.7% (5.5% – 8.0%) and middle-aged adults: *p*_*f*_ = 6.7% (5.3% – 8.4%). The result for the oldest age group, 80 and over: *p*_h_ = 0.0% (0.0% – 3.4%), was substantially lower than any other adult or adolescent age group.

Our results also exhibit age differences in directional household transmission probabilities (HSARs) at given household sizes. The highest estimated average HSAR among all the 9 possible directional age group pairings in this model was from middle-aged and older adults (age 45+) to other middle-aged and older adults: *p*_uu2_ = 68% (56% – 77%) in size-2 households. The lowest estimate for size-2 households was from middle-aged and older adults to children, adolescents, and young adults (age 0-24): *p*_uw2_ = 34% (11% – 53%). We also found significantly smaller acquisition probability for those living in larger households versus smaller ones: *σ*_34_ = 0.43 (0.33 – 0.57), and *σ*_5+_ = 0.22 (0.16 – 0.29). This means that our model estimates that each age-to-age HSAR estimate for size-2 households (Table 2) is lower for size 3-4 households and lower still for size 5+ households. For example, our MLE HSAR estimates among age 45+ adults living together in the three different household size groups are *p*_uu2_ = 68%, *p*_uu34_ = 43%, and *p*_uu5+_ = 31%.

We found age-based differences in the probability that individuals with prior infections tested positive during their infection (as reported on our survey). Infants and preschoolers (age 0–5) had the lowest probability estimate: *ϕ*_*Sa*_ = 16% (6% – 33%), with a confidence interval that overlapped only with that of children aged 6–12: *ϕ*_*Sb*_ = 36% (23% – 49%), reflective of lower infection ascertainment for young children. The MLE for each other age group was above 50%, with middle aged and older individuals having the highest estimates at 73% or higher. The false-positive rate estimate for survey-reported prior test results among individuals with no prior infection was 0.8%: π_*S*_ = 99.2% (98.9% – 99.4%).

We also estimated sensitivity and specificity parameters for each of the antibody tests. Our sensitivity estimates were substantially different for the three tests, with Euroimmun highest at *ϕ*_*AE*_ = 99.2% (97.5% – 99.98%) followed by Siemens, *ϕ*_*AS*_ = 87% (83% – 90%), and Abbott, *ϕ*_*AA*_ = 72% (68% – 76%). Our sensitivity estimate for Euroimmun was higher than what the test manufacturer found from samples of individuals who had tested PCR positive 14+ days earlier. By contrast, our sensitivity estimates for Siemens and Abbott were lower than the respective test manufacturer estimates.

For antibody test specificity, our results produced the lowest estimate for Euroimmun, π_*AE*_ = 98.1% (97.5% – 98.6%) and substantially higher estimates for Abbott, π_*AA*_ = 99.7% (99.4% – 99.8%) and Siemens, π_*AS*_ = 99.85%

(99.65% – 99.95%). Those specificity intervals overlap those obtained by each test manufacturer from testing stored, presumed negative samples collected prior to the COVID-19 pandemic: 99.0% (98.4% – 99.4%) for Euroimmun [22], 99.6% (99.1% – 99.9%) for Abbott [23], and 99.89% (99.66% – 99.98%) for Siemens [24].

#### 3.2.3 Equal contact and reciprocal transmission models

The “equal contact” model, in which the assumptions *σ*_uv_ = *σ*_vv_ = *σ*_wv_ and *σ*_uw_ = *σ*_vw_ = *σ*_ww_ are enforced, produces a MLE result (Table 3) in which the oldest transmission age group has the highest transmissibility, the middle transmission age group has the lowest transmissibility and the highest susceptibility, and the youngest age group has the lowest susceptibility. However, the maximal likelihood of this estimate is much poorer than that of the full model, such that the equal contact model can be rejected by the likelihood ratio test (P = 0.01) and has a poorer AIC score (difference of 5.0) despite using 3 fewer parameters than the full model. By contrast, the “reciprocal transmission” model, in which the assumptions *p*_uv_ = *p*_vu_, *p*_uw_ = *p*_wu_, and *p*_vw_ = *p*_wv_ are enforced, cannot be rejected by the likelihood ratio test and produces a superior AIC score (difference of 4.2) to the full model (Table 3) due to using 3 fewer parameters while achieving a similar likelihood at the MLE.

**Table 3.**
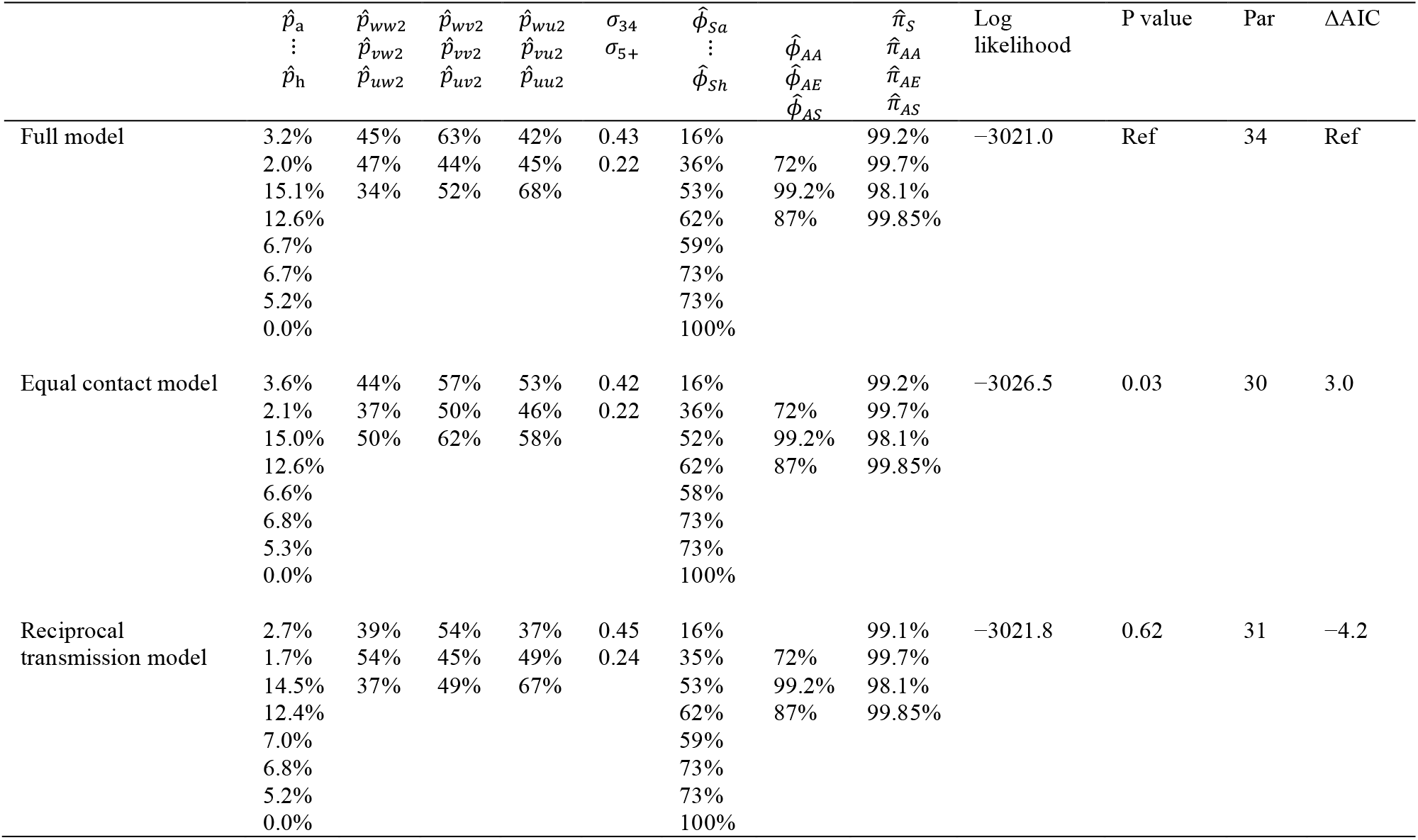
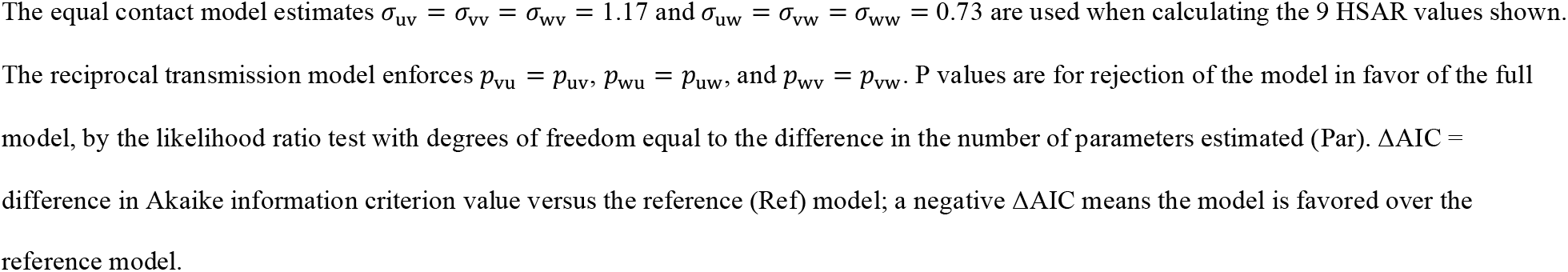
Comparison of equal contact and reciprocal transmission models to full model.

#### 3.2.4 Most parsimonious model results

We tested a series of models using simplifying assumptions determining the age-specific HSAR values, using fewer free parameters to determine the 9 values. The models enforcing equal HSAR to and/or from age group V (age 25-44) produced better AIC scores than those that did not (Table F in Supporting Information). When combining this with other favored assumptions, the best AIC score was produced by a 5-parameter model that used 3 HSAR values to parameterize the 3-by-3 matrix of directional HSAR by age group for a particular household size group, plus the 2 size-effect parameters *σ*_34_ and *σ*_5+_ (Table F in Supporting Information). This model retains the assumption of equal AR between age group V (age 25-44) and all age groups in either direction, and also equal HSAR between age group W (age 0-24) and age groups W and U (age 45+) in either direction, which leaves the HSAR among age group U as the third independent value for a particular household size group.

We also tested simpler models that used two rather than three transmission age groups by consolidating adjacent groups of our three-group model (Table F in Supporting Information). The most parsimonious among those two-group models used ages 0-44 and 45+ as the two groups, with reciprocal transmission – it produced a likelihood inferior to the best three-age-group model just described, with the same number of estimated parameters. Our testing also included even simpler models that used fewer than 5 transmission parameters (Table G in Supporting Information), which could each be rejected in favor of the most parsimonious model (P ≤ 0.03). Those rejected models include models with no age-difference assumptions on transmission (household size being the only correlate with HSAR) or no household size effect (*σ*_34_ = *σ*_5+_ = 1). The simplest model with no age-or size-structure produced a single HSAR estimate of 27% applying to all ages and household sizes, but that result could be strongly rejected (P < 0.0001).

We tested whether adjacent age groups could be consolidated to use fewer free parameters for community acquisition and case ascertainment while achieving comparable model fit. Guided by comparing confidence intervals from the full model MLE results, we tested a model that combined the age groups for community acquisition to reduce the number of groups from 8 to 4, where the 4 groups are ages 0–12 (*p*_a_ = *p*_*b*_), 13–24 (*p*_*c*_ = *p*_*d*_), 25–79 (*p*_*e*_ = *p*_*f*_ = *p*_g_), and 80+ (*p*_h_). For case ascertainment, we reduced the number of groups from 8 to 3: ages 0–12 (*ϕ*_Sa_ = *ϕ*_S*b*_), 13–44 (*ϕ*_S*c*_ = *ϕ*_S*d*_ = *ϕ*_S*e*_), and 45+ (*ϕ*_S*f*_ = *ϕ*_Sg_ = *ϕ*_Sh_). Compared to the full model, the MLE for the model with those new groupings for both community acquisition and case ascertainment could not be rejected by the likelihood ratio test (P = 0.46) and produced superior AIC score with a difference of 9.2 (Table H in Supporting Information). Models using alternate groupings using the same or fewer numbers of age groups produced lower maximal likelihood and inferior AIC scores.

The model combining all the simplifying assumptions producing the best AIC scores in the above comparisons is the most parsimonious overall model we found, producing the best overall AIC score. The most parsimonious model includes 3 household transmission parameters for age-specific HSAR, 2 for household size-adjustment coefficients for HSAR, 4 age-specific importation parameters, 3 age-specific prior case ascertainment parameters, and 7 age-independent sensitivity and specificity parameters (Table 4). The HSAR values shown apply only between members of 2-person households: 66% HSAR between age 45+ individuals residing together, 47% between age 25-44 individuals and anyone else, and 35% between remaining pairings involving age 0-24 individuals with each other or with age 45+ individuals. For households with 3 or 4 residents, the model adjusts those HSAR estimates down to 49%, 31%, and 21%, respectively, and for households with 5 or more residents, the respective HSAR estimates are 35%, 21%, and 13%.

**Table 4.**
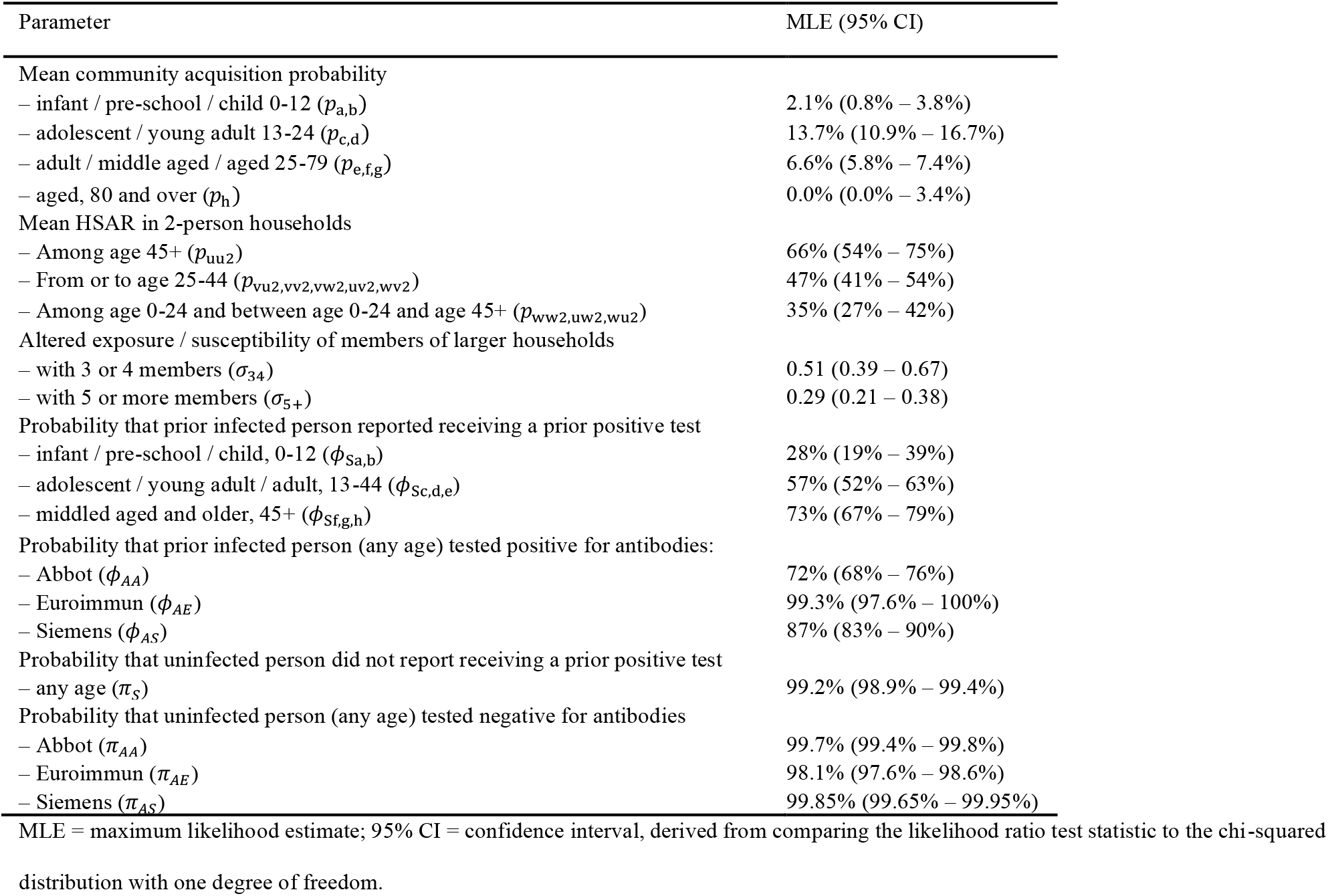
Most parsimonious model with confidence intervals.

| Parameter | MLE (95% CI) |
| --- | --- |
| Mean community acquisition probability |  |
| – infant / pre-school / child 0-12 ($p_{a,b}$) | 2.1% (0.8% – 3.8%) |
| – adolescent / young adult 13-24 ($p_{c,d}$) | 13.7% (10.9% – 16.7%) |
| – adult / middle aged / aged 25-79 ($p_{e,f,g}$) | 6.6% (5.8% – 7.4%) |
| – aged, 80 and over ($p_h$) | 0.0% (0.0% – 3.4%) |
| Mean HSAR in 2-person households |  |
| – Among age 45+ ($p_{uu2}$) | 66% (54% – 75%) |
| – From or to age 25-44 ($p_{vu2,vv2,vw2,uv2,wv2}$) | 47% (41% – 54%) |
| – Among age 0-24 and between age 0-24 and age 45+ ($p_{ww2,uw2,wu2}$) | 35% (27% – 42%) |
| Altered exposure / susceptibility of members of larger households |  |
| – with 3 or 4 members ($\sigma_{3,4}$) | 0.51 (0.39 – 0.67) |
| – with 5 or more members ($\sigma_{5+}$) | 0.29 (0.21 – 0.38) |
| Probability that prior infected person reported receiving a prior positive test |  |
| – infant / pre-school / child, 0-12 ($\phi_{sa,b}$) | 28% (19% – 39%) |
| – adolescent / young adult / adult, 13-44 ($\phi_{sc,d,e}$) | 57% (52% – 63%) |
| – middle aged and older, 45+ ($\phi_{sf,g,h}$) | 73% (67% – 79%) |
| Probability that prior infected person (any age) tested positive for antibodies: |  |
| – Abbot ($\phi_{AA}$) | 72% (68% – 76%) |
| – Euroimmun ($\phi_{AE}$) | 99.3% (97.6% – 100%) |
| – Siemens ($\phi_{AS}$) | 87% (83% – 90%) |
| Probability that uninfected person did not report receiving a prior positive test |  |
| – any age ($\pi_s$) | 99.2% (98.9% – 99.4%) |
| Probability that uninfected person (any age) tested negative for antibodies |  |
| – Abbot ($\pi_{AA}$) | 99.7% (99.4% – 99.8%) |
| – Euroimmun ($\pi_{AE}$) | 98.1% (97.6% – 98.6%) |
| – Siemens ($\pi_{AS}$) | 99.85% (99.65% – 99.95%) |

MLE = maximum likelihood estimate; 95% CI = confidence interval, derived from comparing the likelihood ratio test statistic to the chi-squared distribution with one degree of freedom.

### 3.3 Mechanisms of household transmission differences

The three different age-specific HSARs at a given household size estimated by the most parsimonious model could be explained by a combination of age-based differences in transmissibility, susceptibility, and the amount of household contact. We cannot uniquely estimate the age differences for each of those three mechanisms simultaneously, but we can use our HSAR estimates to constrain their values and explore possible combinations that are consistent with our findings (Fig 1). We showed in section 3.2.3 that assuming an equal amount of household contact between all age groups is inconsistent with our results, as age differences in transmissibility and susceptibility alone cannot reproduce the attack rate matrix. Thus, all depicted possibilities exhibit age-based contact differences (Fig 1). However, uncertainty remains about the potential role of age-based transmissibility and susceptibility differences in combination with different magnitudes of contact differences.

**Fig 1.**
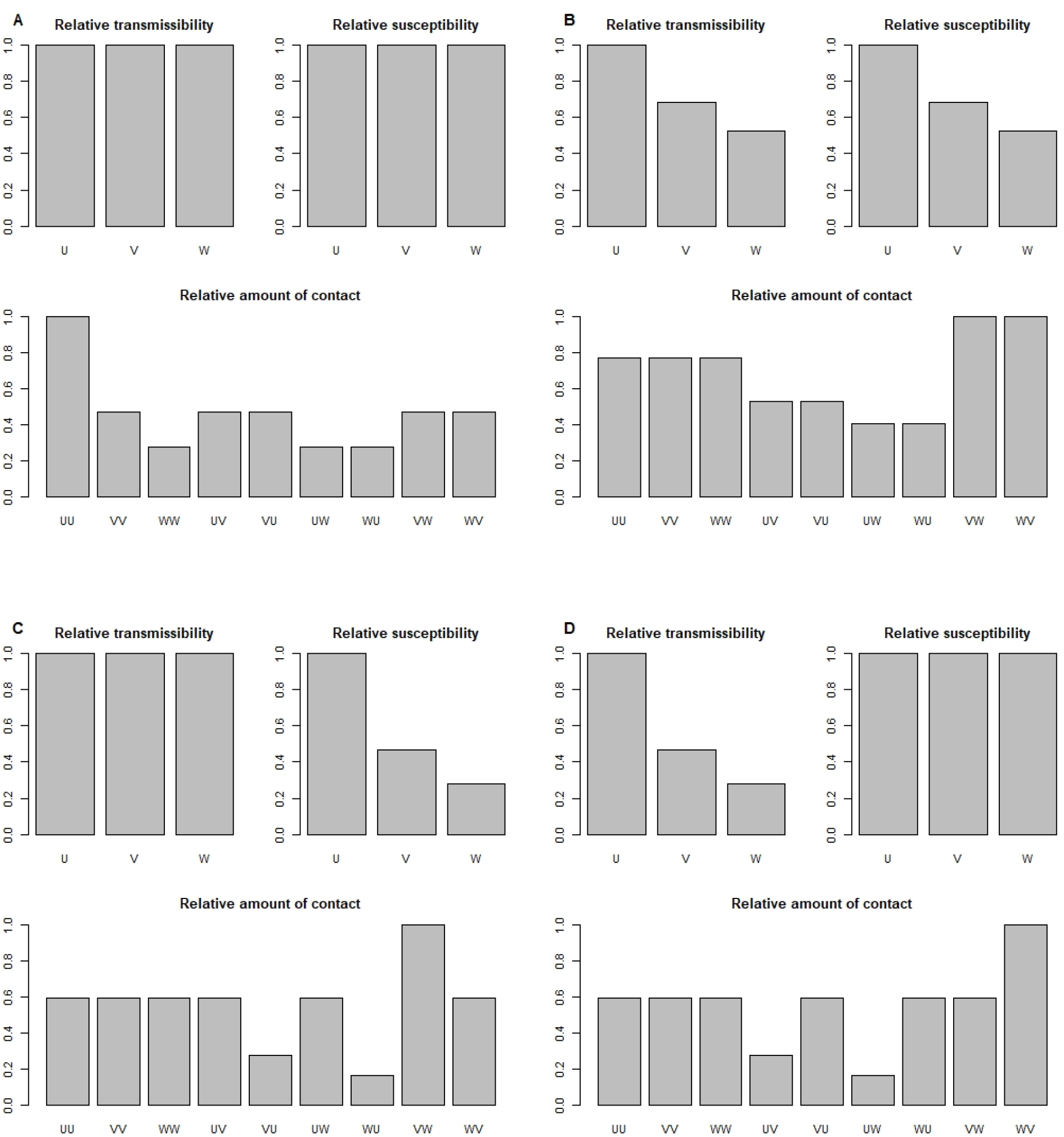
Possible mechanisms of transmission differences consistent with model estimates. W: age group 0-24; V: age group 25-44; U: age group 45+; labels ‘XY’ in relative-amount-of-contact figures refer to the amount of contact between age groups X and Y when X is infected and Y is susceptible. (A) Relative amount of contact when assuming no age difference in susceptibility nor transmissibility; (B) results when assuming equal relative age differences in both susceptibility and transmissibility and no age differences in within-age-group contact; (C) results when assuming no age difference in transmissibility nor in within-age-group contact; (D) results when assuming no age differences in susceptibility nor in within-age-group contact.

One possibility consistent with our results is that transmissibility and susceptibility per contact are independent of age, and that attack rate differences are entirely explained by contact differences (Fig 1A). With this scenario, we estimate the amount of contact between age 25-44 adults and their housemates of any age would have to be about 47% the amount that age 45+ adults contact each other. The amount of contact between age 0-24 and age 0-24 or 45+ would be about 28% the amount of contact within the over-45 age group.

Another possibility is that within-age-group contacts are the same, and the increase of within-group attack rates is explained by both susceptibility and transmissibility increasing with age by equal proportions (Fig 1B). With this scenario, we estimate that susceptibility and transmissibility of those aged 25-44 and 0-24 are about 68% and 53% less than those aged 45+, respectively. This scenario also requires that the highest amount of contact occurred between age 25-44 and age 0-24 individuals.

It is also possible that age groups have equal transmissibility but different susceptibility (Fig 1C) or vice-versa (Fig 1D). However, those scenarios require that contact behavior depends on the age group of the infected person in an infected-susceptible pairing. For example, if aged 25-44 adults are more susceptible than children but equally transmissible, the amount of household contact in a potential transmission scenario between adults and children must be higher when adults were infected and lower when children were infected.

The depicted scenarios illustrate certain boundary cases and are not exhaustive of all possibilities. We derived a full mathematical description of the possible mechanistic parameter combinations consistent with our MLE results (Supporting Information).

### 3.4 Infection risk by age group

Estimates from the most parsimonious model exhibit notable age differences in the overall risk of infection prior to data collection, as well as in relative contribution of non-household and household sources (Fig 2). Members of the youngest (0-12 years) and oldest (80+ years) age groups experienced higher probability of within-household acquisition compared to their risk of acquiring infection from non-household sources. Those in the intermediate age groups experienced most of their acquisition from non-household sources, but within-household sources still contributed between 23% and 41% of their overall infection probability.

**Fig 2.**
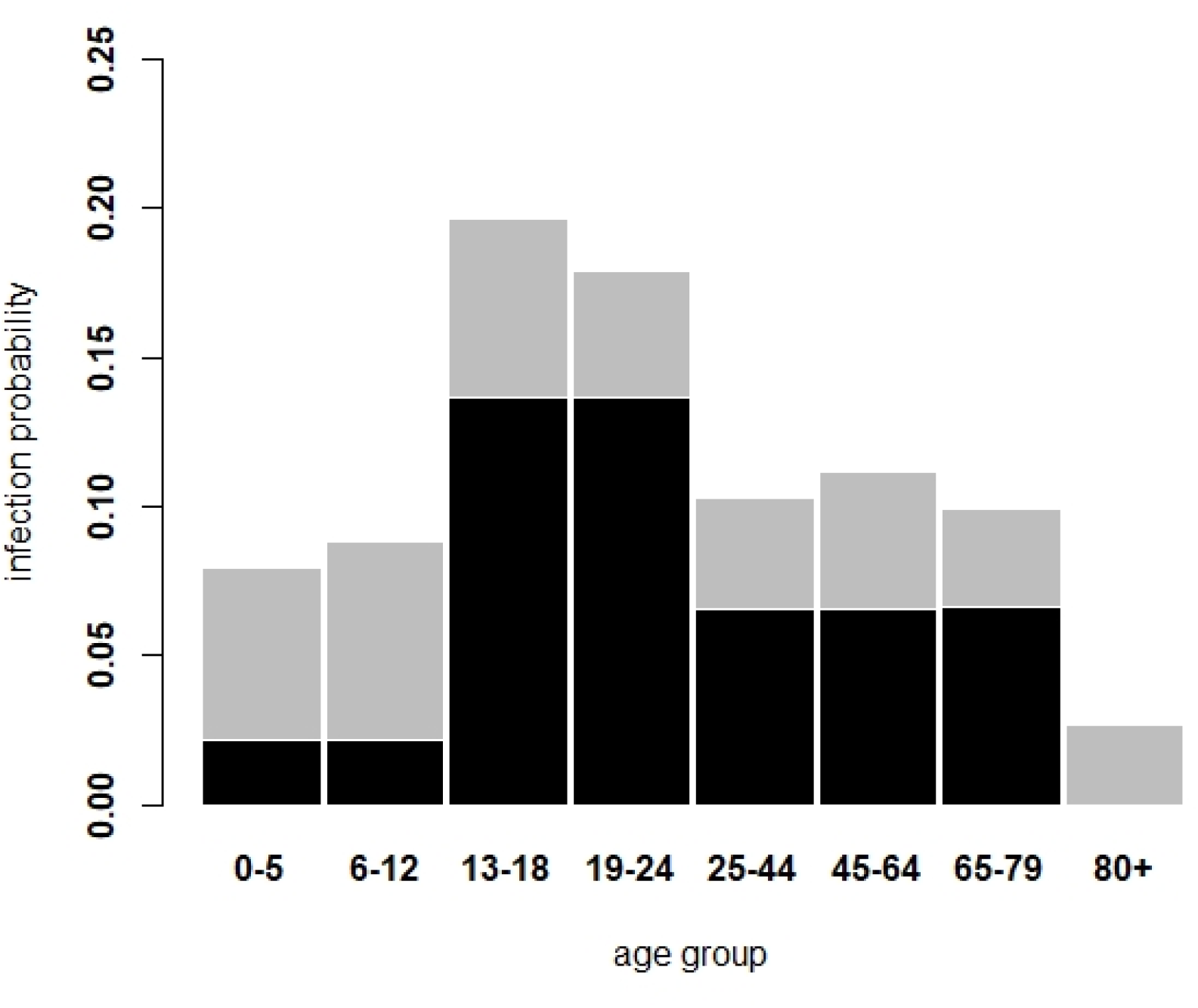
Estimated infection probability of individuals from five age groups from non-household (black) and within-household (grey) sources. Estimates were derived using parameters from the most parsimonious model (Table 4).

Children and adolescents were estimated to have had the highest absolute probability of acquiring infection within the household (6–7%), despite being members of the transmission age group with the lowest HSAR estimates. This was because they lived in larger households than an average adult and were more likely to be exposed to an introduction. Relative to other age groups, adolescents experienced high acquisition risk from both non-household and within-household sources.

The age 25-44 and 45-79 groups experienced similar overall within-household acquisition risk despite the differences in their transmission probabilities; the higher HSAR of age 45+ individuals among themselves was balanced out by their lower rate of acquisition from the youngest age groups. Finally, the age 80+ age group were estimated to have negligible non-household acquisition risk and the lowest within-household risk, the latter due to their living with individuals in the younger groups less frequently.

## 4 Discussion

Our novel mathematical model of household infection, transmission, and test data accuracy can simultaneously incorporate multiple highly important features of SARS-CoV-2: variability in transmissibility of infected people and heterogeneity in non-household acquisition, case ascertainment, and household transmission across age groups and household sizes. Our model could produce nuanced estimates for household transmission dynamics from data and provide novel insights about potential mechanisms of disparate infection rates across households of different sizes and age combinations.

Our findings show strong evidence that adolescents and young adults aged 13-24 were more likely than younger children and older adults to acquire SARS-CoV-2 infection outside the household prior to data collection in early 2021 in Utah. Children aged 12 and younger were less likely to acquire outside the household than any adult group other than those aged 80 and over. We found little evidence of any acquisitions by the age 80+ group from outside the household, suggesting that the primary risk to these elderly individuals living in households may have been transmission from their housemates. Our data did not include elderly residing in congregate living facilities such as nursing homes, where residents likely experienced a much different risk profile compared to aged individuals covered by our sampling of households [29].

The results also suggest that there were differences in household transmission rates across age group pairings. The highest age-specific HSAR for a given household size was from middle-aged and older adults (age 45+) to other middle-aged and older adults. The next-highest HSAR estimate was for adults aged 25-44, from and to all age groups. The lowest HSAR estimate was for those younger than 25 among each other and to and from those aged 45 and older. That age pattern of attack rates was inconsistent with a model that assumes an age-independent rate of contact among infected and susceptible pairs of housemates. With that equal-contact model, the age group most susceptible to acquiring infection for a given amount of infectious contact would experience the highest attack rate from all three age groups. However, our findings showed that, while the 45-and-older group experienced the highest attack rate from age 45+ housemates, they experienced a lower attack rate from the under-25 group compared to that experienced by the 25-44 group. Similarly, while infected 45-and-older individuals exhibited the highest attack rate to other 45-and-older housemates, they transmitted at a lower rate to under-25 housemates compared to the corresponding attack rate from the 25-44 group. Therefore, our findings suggest that contact within households was not uniform across age groups.

Our finding that the attack rate estimates were similar in both directions for each given age-group pairing is consistent with a hypothesis that there are not substantial age differences in susceptibility or transmissibility for a given rate of contact. For example, if 45-and-older individuals were substantially more susceptible to acquiring infection from a given contact compared to those under 25, one might expect the under-25 to over-45 attack rate to be higher than the over-45 to under-25 attack rate. However, it is possible that an equal attack rate in both directions could be explained by multiple mechanistic differences that cancel each other out, such as age-dependent susceptibility, transmissibility, or contact behavior changes during infection.

Additional data on age-structured, transmission-relevant contact behavior in households would help further constrain the possible mechanisms driving the age differences in transmission we found. For example, if data showed that contact between younger parents and younger children (i.e. between “V” and “W” groups in our model) was more substantial than contact between other age pairings within households, and especially if that contact persisted during illness because of the necessities of childcare [30], then our findings would suggest stronger evidence that younger age groups had lower susceptibility and/or transmissibility per contact compared to older age groups (Fig 1 panels B-D).

We also found lower HSARs for larger household sizes, even when controlling for our age difference findings. This finding is consistent with other household transmission studies [11-13] that favored models with transmission rates scaled down with increasing household size, e.g. via frequency-dependent-like transmission assumptions. The precise mechanisms by which transmission probabilities for a given age-to-age pairing might decrease with increasing household size are unclear. There could be contact rate differences among members of larger vs. smaller families [12], or risk differences due to domicile size and layout or demographic factors other than age that correlate with household size. Differences in precaution behavior to prevent transmission might also play a role, particularly in later generations of infection after a given household introduction, which would be a larger factor for more populous households where multiple rounds of transmission are possible.

In combination, our findings for age-dependency and household size-dependency of household transmission result in dramatically HSAR depending on the scenario. Our model’s HSAR estimates range from 66% for middle-aged or older adults living alone together, down to 13% among age 0-24 individuals or between age 0-24 and 45+ individuals living together in households of size 5 or more. This finding suggests that standard estimates of a single averaged HSAR can be highly misleading if used to judge the risk of transmission in all households.

Our estimates quantifying transmission variability are consistent with our prior finding that the low-variability binomial transmission assumption traditionally used in household models can be rejected in favor of models incorporating higher variability (lower value of a dispersion parameter) for individual transmissibility [18]. Notably, our model factors out potential sources of variability that could correlate with age and/or household size. Yet we still found that incorporating additional variability for individual transmissibility within members of the same age/size groups was warranted, suggesting that other sources of variability beyond those determined by age and household size are important to represent [2].

The dispersion parameter, *k*, that we used to quantify transmission variability is analogous to the dispersion parameter *k* commonly used in negative binomial transmission models, in that it derives from the shape parameter of gamma-distributed individual transmissibility [1]. However, in the negative binomial model, *k* represents all potential contributions to variability, including variability in number of people contacted [31] and variation in non-contact-based characteristics that might correlate with age. Our model explicitly represents those features and leaves *k* to represent any remaining contributions to variability. I.e., we use *k* to represent variability of transmission probability to *each* housemate of a given age group, from a given age group, in a household of a given size. Therefore, if contact degree and and/or age/size-based differences contribute significantly to overall transmission variation, we would expect our estimate of *k* to be higher (lower variability) than estimated *k* from a negative binomial model. Indeed, a systematic review [32] found that 93% of 28 studies estimated *k* < 1 for the negative binomial model fit to COVID-19 transmission data, whereas we did not find strong support for *k* < 1 in our model (*k* = 1 could not be rejected and produced the most parsimonious model fit). However, our rejection of the lower variance binomial assumption (*k* → ∞) suggests the possibility that a large portion of overall transmission variability could be attributed to factors other than number of contacts or age/size-associated differences, such as the duration of infectiousness, organism shedding rate [33], or individual contact timing and intensity.

Our detailed data including results from up to three different antibody tests also presented a unique opportunity for comparing test accuracies against each other and against data from the test manufacturers or other sources. Our independent estimates of the antibody test specificities were remarkably consistent with results from independent data. However, our results for antibody test sensitivities included some disagreements with other sources, particularly for the Euroimmun test. Further research could illuminate why our Euroimmun sensitivity estimates were so high, perhaps by building a model that utilizes the quantitative Euroimmun results to investigate whether the manufacturer-suggested positive/indeterminate/negative thresholds were less appropriate for our implementation.

Our extensive antibody test data also allowed us to estimate case ascertainment across age groups. By comparing survey data about whether antibody-positive individuals had ever tested positive prior to serological testing, we found a high level of age dependence: younger individuals with a prior infection were substantially less likely to have tested positive during their infection compared to older age groups. This finding provides evidence that case ascertainment in Utah had been substantially lower for children aged 0-12 compared to adolescents and adults, and somewhat lower for those aged 13-44 compared to those middle-aged and older, perhaps suggesting that those younger groups were less likely to experience symptoms that warranted seeking testing [34].

This study has several limitations. We did not test factors other than age composition and household size that might affect the way community acquisition risk or within-household transmission varies by household. Some households could have experienced higher collective community acquisition risk than others of similar age composition due to living in higher-risk locations, common work or school exposure, or common social exposure. Household transmission could also be driven by non-age-based properties of households such as age-independent contact behavior, health composition of household members, physical properties of the domicile such as size and ventilation, or other properties that could increase infection or transmission risk of all household members together. It is possible that further expansion of our model to include correlates with non-age data and/or household-level variability distributions for community acquisition or household transmission could provide further insights, but we suspect that these additions would push the limits of parameter identifiability against our data.

Some members of participating households might have been actively or very recently infected with SARS-CoV-2 at the time of sampling for serological testing. When ongoing or recent household outbreaks involved multiple infections, some of the infected members could have tested seropositive while others had yet to become infected or had yet to seroconvert on the day of sampling. For that scenario, our assumption that household outbreaks had reached their final size would be violated, contributing to underestimates of HSAR. We suspect that the proportion of those households was low, given the short time window in which those conditions would apply in a multi-person outbreak, and households might have been unlikely to choose to travel to a testing site while experiencing or recovering from symptomatic infection.

Our findings may contain biases related to non-participation rates of households that were selected and approached for inclusion in the study. Our data collection in the school district testing phase included a complicated sampling design with weights introduced partly to account for different rates of nonresponse across demographic strata. For simplicity we ignored these details and sampling weights for the analysis presented here. In the hotspot testing phase households were selected for invitation by simple random sample of addresses, and those choosing to participate may not be representative of those areas. Also, while the included sampling phases and locations cover a broad area of the most populous region in Utah, there may be important differences in households both within those areas and from the areas of Utah not covered. Thus, households with higher COVID-19 risk may be overrepresented or underrepresented in our data relative to their frequency in the broader population of households in Utah.

We used participants’ prior vaccination information only to indicate whether positive test results for the two spike-protein-based antibody tests might have been triggered by the vaccine rather than prior SARS-CoV-2 infection; we did not attempt to assess whether the vaccine produced a protective affect against community and/or within-household acquisition in the period after receiving one or two vaccine doses. Our raw data show that vaccinated individuals tested positive by the Abbott (nucleocapsid) test at a lower rate than non-vaccinated individuals, particularly in the 65-79 age group, which had received higher vaccination coverage than younger age groups. Thus, it is possible that including a vaccination effect parameter on community acquisition or household transmission probability would produce a result suggesting a reduced infection probability. However, such a result would not be conclusive of a causal effect of the vaccine on acquisition risk, for the following reasons.

As vaccines had only recently become available in early 2021, only 12% of participants had received the full two-dose series prior to serological sampling, and about half of those had received their second dose less than one month prior. Thus, even the minority of those participants who were considered fully vaccinated at the time of testing had spent only a small portion of their cumulative time at risk since the onset of the pandemic in a potentially protected state. The subset of participants who had experienced both prior infection and vaccination could include both those who were infected prior to vaccination as well as break-through infections post vaccination, and inferring the relative timing of these events would be difficult with our data. Also, inferring a potential causal effect of vaccination on infection risk would be hampered by confounders, including risk-based vaccination eligibility, the potential reverse causal effect of known prior infection on the decision to vaccinate, and potential correlations between exposure risk and vaccine access and/or refusal among those eligible. If the vaccinated participants did experience substantial COVID-19 exposures between their vaccination and serological testing, and the vaccine substantially reduced their likelihood of infection by those exposures, then our results for the relative acquisition probabilities of older age groups (who had higher vaccine coverage) might be biased low.

Our dataset’s collection period ended on April 10, 2021, just prior to the widespread emergence of the Delta variant of SARS-CoV-2, which was first detected in Utah in mid-April 2021 and spread rapidly in the following weeks [35]. Thus, our estimates may not accurately reflect infection and transmission characteristics of dominant SARS-CoV-2 strains circulating in the U.S. and elsewhere from late Spring 2021 to the present, particularly because different strains have been found to exhibit different household transmission rates [36]. Also, as the state of Utah has a younger population, larger average household size, and other demographic differences compared to U.S. national averages [18], our household-based epidemiological estimates might be atypical. However, as we expressed our household transmission estimates as per-capita transmission probabilities from and to particular age groups and household sizes, our findings could serve as tool to study how overall household transmission patterns would differ in communities with different household size and age distributions.

In conclusion, we found evidence of important age differences in COVID-19 epidemiology in Utah households during 2020 and early 2021, including differences in case ascertainment, non-household acquisition, and within-household transmission. Household transmission differences by age occurred in patterns that suggest substantial differences in household contact rates between different age groups. A better understanding of those differences might be crucial to deciphering community-wide transmission patterns and mitigation efforts.

## Data Availability

The data and computer codes used to produce the results in the present study are publicly available at https://github.com/damontoth/householdTransmission2021.

## 5 Acknowledgments

The authors acknowledge David Swerdlow, Fred Angulo, Jessica Atwell, and Farid Khan for inspiration for this data modeling work and comments on this manuscript. We acknowledge Stephen Alder, Adam Looney, Tom Greene, Brian Orleans, Yue Zhang, and other members of the Utah HERO Project team and the University of Utah Study Design & Biostatistics Center who helped design and/or implement the household participant recruitment and survey effort. We acknowledge Julio Delgado for directing the validation and implementation of the antibody assays at ARUP Laboratories and overseeing the validation of finger prick sample collection, as well as the staff at ARUP Laboratories who processed and tested sample specimens to produce the antibody data used in our analysis.

## Supporting Information

### Participant recruitment

Participants contributing to our dataset were recruited during two different phases of the Utah HERO Project. The first phase targeted households within the geographic boundaries of two school districts (“school district testing”): the Salt Lake City School District, identical to the boundaries of Salt Lake City, and the adjacent Granite School District, which includes other urban areas in Salt Lake County. These geographies were chosen to enable analysis of relationships between school policies [21] and community transmission of SARS-CoV-2 and to understand the extent of infection in this urban population.

For the school district testing phase, participants were recruited and enrolled starting from January 7, 2021, and samples were collected from January 14 to April 10, 2021. The source population consisted of all households in the two school district boundaries as defined in 2020-2021, for a total of 83,353 and 138,397 possible households in Salt Lake City and Granite School Districts, respectively. Households were chosen using a random sampling design as previously described [20]. A total of 75,067 households were invited by mail to participate, of which 35,936 were in the Salt Lake City School District and 39,131 were in the Granite School District.

The second phase of recruitment contributing to our dataset occurred in response to a spike in COVID-19 cases in specific areas (“hotspot testing”). The Utah Department of Health had notified the Utah HERO Project team of an especially high incidence of COVID-19 cases in early 2021 among communities in the southern portion of Salt Lake County and the northern portion of Utah County. In response, the HERO Project conducted a simple random sample of households in the following cities/towns in Salt Lake County (Draper, Highland, South Jordan, West Jordan) and Utah County (Alpine, American Fork, Bluffdale, Cedar Hills, Eagle Mountain, Herriman, Lehi, Lindon, Pleasant Grove, Riverton, Saratoga Springs) in the State of Utah. Households that had been tested during previous phases of the Utah HERO Project were not eligible for participation in the hotspot testing phase. A total of 21,480 households were invited by mail to participate: 10,601 in Salt Lake County and 10,879 in Utah County. Recruitment for hotspot testing began in late February 2021, and samples were collected from March 2 to March 13, 2021.

For both the school district and hotspot testing phases of recruitment, the Utah HERO Project team sent a postcard and letter to each household in the sampling frame encouraging household members to participate. Study participants received testing free of charge, and each household member who completed the survey and provided a blood sample received a $30 gift card.

### Derivation of household transmission distribution F_yz_

When an infected household member has susceptible housemates, we require the probabilities that the infected person transmits to given numbers of them. The susceptible housemates can be members of up to three different age groups: U, V, and W. We assume the probability of transmission to a susceptible group-u housemate is 1 *− e*^*−x*^, where *x* is a gamma-distributed random variable with shape *k* and rate *r*. Then the transmission probability to a susceptible group-v housemate is 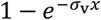, and to a susceptible group-w housemate is 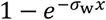, where and *σ*_v_ and *σ*_w_ are constants governing the altered transmission probability to age groups V and W relative to age group U.

***F***_**yz**_ is the joint probability of the infected person directly transmits to **y** = (*y*_u_, *y*_v_, *y*_w_) of **z** = (z_u_, z_v_, z_w_) susceptible housemates. We show here that these assumptions lead to the formula for this probability stated in the main text.

We use the binomial distribution with individual success probability distributed as described above:

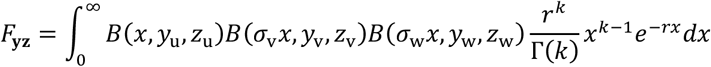

Where *B* is the probability mass function of the binomial distribution taking the following form:

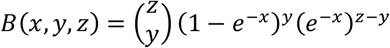

The expression for *B* can be rewritten by converting (1 *− e*^*−x*^)^*y*^ into a sum:

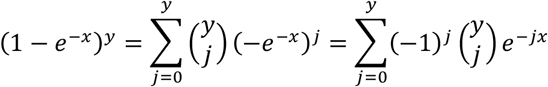

So

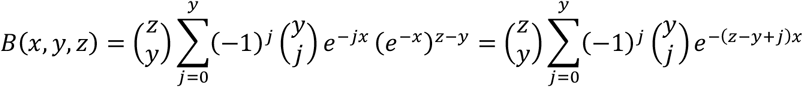

Substituting within the integral for F we get:

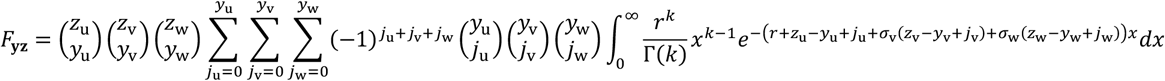

The integral can be solved:

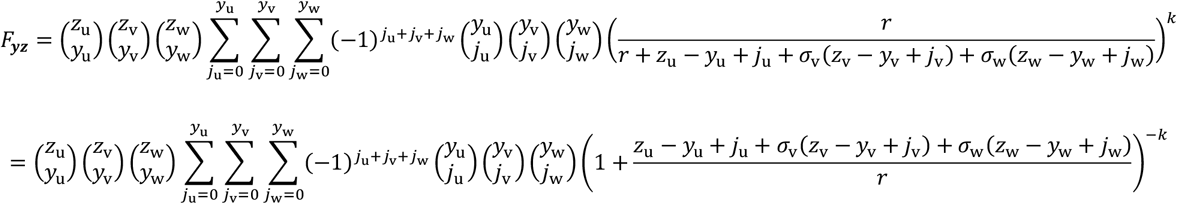

### Mathematical description of mechanisms of household transmission differences

In our model with exponentially distributed transmissibility, the average household secondary attack rate *P*_*XY*_ from an infected person in age group X to a susceptible housemate in age group Y can be expressed:

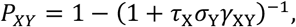

where τ_X_ is the transmissibility of a person in age group X, *σ*_Y_ is the susceptibility of a person in age group Y, and *γ*_XY_ is the amount of contact between people in age groups X and Y when the age-group-X person is infected and the age-group-Y person is susceptible to infection. More specifically, we can define τ as the average total number of organisms shed over the period of infection, *σ* as the per-organism probability of infection given a dose of exposure (as in the exponential dose-response model), and *γ* as the portion of total shed organisms to which the susceptible contact is exposed.

Our MLE results provide *P*_*XY*_ for each of the nine directional attack rates among the three age-groups in our model. Because there are 15 unknown values in the above formulation (3 τ_X_ values, 3 *σ*_Y_ values and 9 *γ*_XY_ values), there is not a unique solution for all values given the 9 equations. We require 6 additional equation-based assumptions to arrive at a unique solution. First, because we have no independent information about absolute transmissibility (count of shed organisms), susceptibility (dose-response), or contact exposures in households, we first make an arbitrary assumption for these values for a particular age pairing, constrained by its MLE attack rate, and write equations for the ratios of each component for the other age group against the chosen index. Using U-U as the baseline, we reduce to the following set of 8 equations:

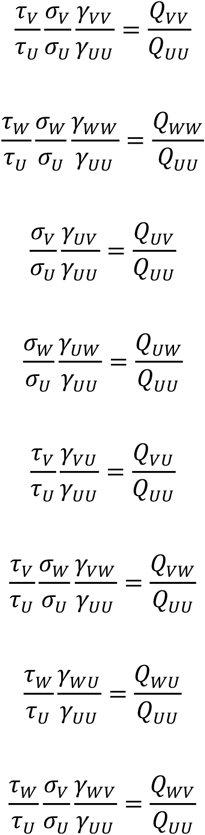

where *Q*_*XY*_ = (1 *− P*_*XY*_)^*−*1^ *−* 1. In these 8 equations, there are 12 unknown ratios: two each for τ and *σ* values for groups V and W relative to U and 8 for *γ* values relative to the U-U contact assumption. Note that the unknown ratios are not dependent on the values of τ_*U*_, *σ*_*U*_, and *γ*_*UU*_, so the assumptions for their values are arbitrary if our goal is to explore the ratios.

We need 4 additional assumptions to determine potential unique solutions for the 12 unknown ratios in the above equation. In the main text, we made the following sets of assumptions to illustrate a range of possibilities, corresponding to Figure 2A-D (main text):

A:

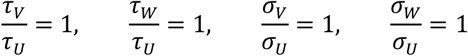

B:

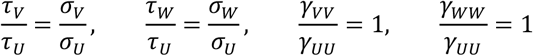

C:

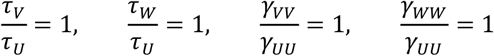

D:

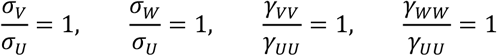

**Table A.** Household recruitment and number of individuals tested by phase and location.

| Phase / Location | Households contacted | Households tested (response %) | Individuals tested | Individuals per household |
| --- | --- | --- | --- | --- |
| School district: Granite | 39,131 | 1,123 (2.9%) | 2,539 | 2.26 |
| School district: Salt Lake City | 35,936 | 1,677 (4.7%) | 3,262 | 1.94 |
| Hotspot: Salt Lake County | 10,601 | 290 (2.7%) | 907 | 3.12 |
| Hotspot: Utah County | 10,879 | 291 (2.7%) | 999 | 3.43 |
| **Total** | **96,547** | **3,381 (3.5%)** | **7,707** | **2.28** |

**Table B.** Positivity rates by age and vaccination status.

| MeSH age category | Prior positive / total participants | Abbott positive / total determinate | Euroimmun positive / total determinate | Siemens positive / total determinate |
| --- | --- | --- | --- | --- |
| All ages | Unvax: 349 / 4518 (7.7%)<br>Vax1: 37 / 610 (6.1%)<br>Vax2: 41 / 727 (5.6%) | Unvax: 328 / 3436 (9.5%)<br>Vax1: 37 / 598 (6.2%)<br>Vax2: 40 / 712 (5.6%) | Unvax: 505 / 3587 (14.2%)<br>Vax1: 378 / 513 (73.7%)<br>Vax2: 664 / 672 (98.8%) | Unvax: 392 / 3434 (11.4%)<br>Vax1: 373 / 597 (62.5%)<br>Vax2: 693 / 713 (97.2%) |
| Newborn/ Infant/ Preschool (0 – 5) | Unvax: 7 / 350 (2.0%) | Unvax: 0 / 2 (0%) | Unvax: 32 / 312 (10.3%) | Unvax: 0 / 1 (0%) |
| Child (6 – 12) | Unvax: 21 / 633 (3.3%) | Unvax: 0 / 8 (0%) | Unvax: 55 / 573 (9.6%) | Unvax: 0 / 8 (0%) |
| Adolescent (13 – 18) | Unvax: 47 / 394 (11.6%)<br>Vax1: 0 / 8 (0%)<br>Vax2: 0 / 3 (0%) | Unvax: 57 / 369 (15.4%)<br>Vax1: 0 / 8 (0%)<br>Vax2: 0 / 3 (0%) | Unvax: 76 / 322 (23.6%)<br>Vax1: 4 / 7 (57.1%)<br>Vax2: 3 / 3 (100%) | Unvax: 72 / 369 (19.5%)<br>Vax1: 5 / 8 (62.5%)<br>Vax2: 3 / 3 (100%) |
| Young adult (19 – 24) | Unvax: 26 / 247 (10.5%)<br>Vax1: 3 / 14 (21.4%)<br>Vax2: 2 / 27 (7.4%) | Unvax: 26 / 233 (11.1%)<br>Vax1: 3 / 13 (23.1%)<br>Vax2: 2 / 27 (7.4%) | Unvax: 39 / 189 (20.6%)<br>Vax1: 9 / 12 (75.0%)<br>Vax2: 25 / 25 (100%) | Unvax: 33 / 233 (14.2%)<br>Vax1: 10 / 13 (76.9%)<br>Vax2: 27 / 27 (100%) |
| Adult (25 – 44) | Unvax: 115 / 1552 (7.4%)<br>Vax1: 12 / 178 (6.7%)<br>Vax2: 17 / 242 (7.0%) | Unvax: 120 / 1510 (7.9%)<br>Vax1: 11 / 176 (6.3%)<br>Vax2: 15 / 236 (6.4%) | Unvax: 157 / 1191 (13.2%)<br>Vax1: 116 / 153 (75.8%)<br>Vax2: 214 / 215 (99.5%) | Unvax: 141 / 1510 (9.3%)<br>Vax1: 121 / 176 (68.8%)<br>Vax2: 235 / 236 (99.6%) |
| Middle aged (45 – 64) | Unvax: 98 / 931 (10.5%)<br>Vax1: 10 / 236 (4.2%)<br>Vax2: 13 / 164 (7.9%) | Unvax: 86 / 910 (9.5%)<br>Vax1: 12 / 232 (5.2%)<br>Vax2: 16 / 160 (10.0%) | Unvax: 107 / 710 (15.1%)<br>Vax1: 143 / 209 (68.4%)<br>Vax2: 140 / 145 (96.6%) | Unvax: 100 / 909 (11.0%)<br>Vax1: 132 / 231 (57.1%)<br>Vax2: 152 / 160 (95.0%) |
| Aged (65 – 79) | Unvax: 34 / 381 (8.9%)<br>Vax1: 11 / 160 (6.9%)<br>Vax2: 9 / 251 (3.6%) | Unvax: 39 / 375 (10.4%)<br>Vax1: 10 / 155 (6.5%)<br>Vax2: 7 / 247 (2.8%) | Unvax: 41 / 273 (15.0%)<br>Vax1: 99 / 123 (80.5%)<br>Vax2: 243 / 245 (99.2%) | Unvax: 45 / 375 (12.0%)<br>Vax1: 99 / 155 (63.9%)<br>Vax2: 240 / 248 (96.8%) |
| Aged (80 and over) | Unvax: 1 / 30 (3.3%)<br>Vax1: 1 / 14 (7.1%)<br>Vax2: 0 / 40 (0%) | Unvax: 0 / 29 (0%)<br>Vax1: 1 / 14 (7.1%)<br>Vax2: 0 / 39 (0%) | Unvax: 1 / 17 (5.9%)<br>Vax1: 7 / 9 (77.8%)<br>Vax2: 39 / 39 (100%) | Unvax: 1 / 29 (3.4%)<br>Vax1: 6 / 14 (42.9%)<br>Vax2: 36 / 39 (92.3%) |

MeSH = Medical Subject Headings [28]; Unvax = unvaccinated (participants who reported receiving no prior vaccine doses); Vax1 = participants
who reported receiving exactly 1 vaccine dose; Vax2 = participants who reported receiving 2 vaccine doses; total determinate = total number of
individuals receiving a test result of positive or negative; prior positive = individuals who reported on the survey that they had tested positive for
COVID-19

**Table C.**
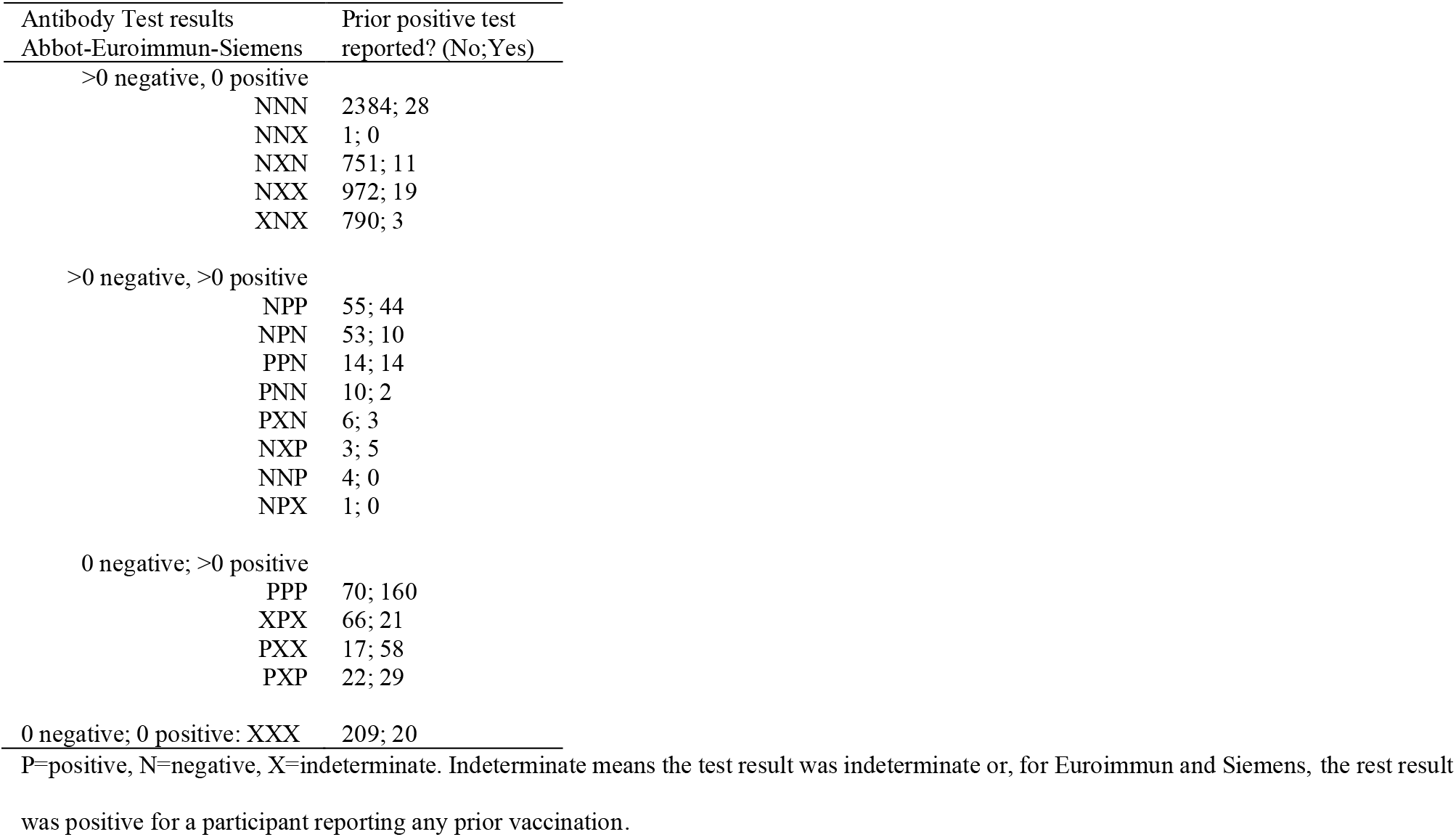
Frequency of test and survey result combinations among individuals in the data.

**Table D.** Transmission variability model comparison.

| | $\hat{p}_a$<br>:<br>$\hat{p}_h$ | $\hat{p}_{ww2}$<br>$\hat{p}_{vw2}$<br>$\hat{p}_{uw2}$ | $\hat{p}_{wv2}$<br>$\hat{p}_{vv2}$<br>$\hat{p}_{uv2}$ | $\hat{p}_{wu2}$<br>$\hat{p}_{vu2}$<br>$\hat{p}_{uu2}$ | $\sigma_{34}$<br>$\sigma_{5+}$ | $\hat{k}_w$<br>$\hat{k}_v$<br>$\hat{k}_u$ | $\hat{\phi}_{sa}$<br>:<br>$\hat{\phi}_{sh}$ | $\hat{\phi}_{AA}$<br>$\hat{\phi}_{AE}$<br>$\hat{\phi}_{AS}$ | $\hat{\pi}_s$<br>$\hat{\pi}_{AA}$<br>$\hat{\pi}_{AE}$<br>$\hat{\pi}_{AS}$ | Log<br>likelihood | P value | Par | $\Delta AIC$ |
| --- | --- | --- | --- | --- | --- | --- | --- | --- | --- | --- | --- | --- | --- |
| Gamma-distributed<br>transmissibility | 3.2%<br>2.0%<br>15.0%<br>12.6%<br>6.6%<br>6.7%<br>5.2%<br>0.0% | 47%<br>48%<br>34% | 68%<br>44%<br>51% | 42%<br>44%<br>68% | 0.42<br>0.22 | 0.75<br>1.00<br>1.82 | 16%<br>35%<br>53%<br>62%<br>59%<br>73%<br>73%<br>100% | 72%<br>99.2%<br>87% | 99.1%<br>99.7%<br>98.1%<br>99.85% | -3020.8 | Ref | 37 | Ref |
| Exponentially<br>distributed<br>transmissibility | 3.2%<br>2.0%<br>15.1%<br>12.6%<br>6.7%<br>6.7%<br>5.2%<br>0.0% | 45%<br>47%<br>34% | 63%<br>44%<br>52% | 42%<br>45%<br>68% | 0.43<br>0.22 | 1<br>1<br>1 | 16%<br>36%<br>53%<br>62%<br>59%<br>73%<br>73%<br>100% | 72%<br>99.2%<br>87% | 99.2%<br>99.7%<br>98.1%<br>99.85% | -3021.0 | 0.95 | 34 | -5.7 |
| No-variance<br>transmissibility | 2.7%<br>1.9%<br>14.9%<br>12.5%<br>6.6%<br>6.8%<br>5.2%<br>0.0% | 46%<br>45%<br>35% | 69%<br>42%<br>52% | 37%<br>43%<br>69% | 0.51<br>0.27 | ∞<br>∞<br>∞ | 17%<br>36%<br>53%<br>62%<br>58%<br>73%<br>73%<br>100% | 72%<br>99.2%<br>87% | 99.1%<br>99.7%<br>98.1%<br>99.85% | -3022.7 | 0.28 | 34 | -2.2 |

P values are for rejection of the model in favor of the full model, by the likelihood ratio test with degrees of freedom equal to the difference in number of parameters (Par). Par = number of independent parameters simultaneously estimated. $\Delta AIC$ = difference in Akaike information criterion value versus the reference (Ref) model; a negative $\Delta AIC$ means the model is favored over the reference model, and the lowest $\Delta AIC$ model is favored over all the others.

**Table E.**
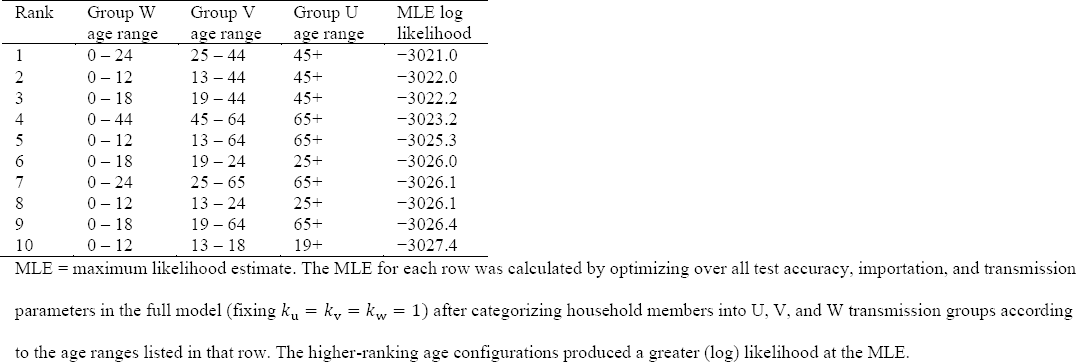
Maximum likelihood results for different transmission age groupings.

**Table F.** Comparison of simpler age- and size-structured transmission models to full model.

| | $\hat{p}_{ww2}$<br>$\hat{p}_{vw2}$<br>$\hat{p}_{uw2}$ | $\hat{p}_{wv2}$<br>$\hat{p}_{vv2}$<br>$\hat{p}_{uv2}$ | $\hat{p}_{wu2}$<br>$\hat{p}_{vu2}$<br>$\hat{p}_{uu2}$ | $\sigma_{34}$<br>$\sigma_{5+}$ | Log likelihood | P value | Par | ΔAIC |
| --- | --- | --- | --- | --- | --- | --- | --- | --- |
| Full model | 45%<br>47%<br>34% | 63%<br>44%<br>52% | 42%<br>45%<br>68% | 0.43<br>0.22 | -3020.98 | Ref | 11 | Ref |
| Equal HSAR from U | 50%<br>56%<br>**62%** | 71%<br>46%<br>**62%** | 49%<br>49%<br>**62%** | 0.32<br>0.16 | -3025.25 | 0.01 | 9 | 4.5 |
| Equal HSAR from V | 45%<br>**45%**<br>32% | 62%<br>**45%**<br>51% | 42%<br>**45%**<br>67% | 0.44<br>0.21 | -3021.03 | 0.95 | 9 | -3.9 |
| Equal HSAR from W | **47%**<br>46%<br>28% | **47%**<br>46%<br>52% | **47%**<br>38%<br>66% | 0.48<br>0.24 | -3023.33 | 0.10 | 9 | 0.7 |
| Equal HSAR to U | 53%<br>51%<br>30% | 69%<br>45%<br>52% | **59%**<br>**59%**<br>**59%** | 0.35<br>0.17 | -3023.98 | 0.05 | 9 | 2.0 |
| Equal HSAR to V | 38%<br>42%<br>29% | **48%**<br>**48%**<br>**48%** | 34%<br>36%<br>65% | 0.56<br>0.30 | -3021.83 | 0.43 | 9 | -2.3 |
| Equal HSAR to W | **39%**<br>**39%**<br>**39%** | 57%<br>42%<br>51% | 32%<br>36%<br>65% | 0.53<br>0.27 | -3021.36 | 0.68 | 9 | -3.2 |
| Equal HSAR to and from V | 37%<br>**47%**<br>33% | **47%**<br>**47%**<br>**47%** | 34%<br>**47%**<br>66% | 0.51<br>0.28 | -3022.17 | 0.88 | 7 | -5.6 |
| Consolidate W and V | **47%**<br>**47%**<br>37% | **47%**<br>**47%**<br>37% | **39%**<br>**39%**<br>67% | 0.48<br>0.24 | -3023.64 | 0.37 | 6 | -4.7 |
| Consolidate V and U | 47%<br>**43%**<br>**43%** | 54%<br>**56%**<br>**56%** | 54%<br>**56%**<br>**56%** | 0.41<br>0.21 | -3027.18 | 0.03 | 6 | 2.4 |
| Consolidate W and V;<br>reciprocal transmission | **48%**<br>**48%**<br>40% | **48%**<br>**48%**<br>40% | 40%<br>40%<br>67% | 0.46<br>0.22 | -3023.69 | 0.49 | 5 | -6.6 |
| Equal HSAR to and from V;<br>equal HSAR W-to-W, U-to-W, & W-to-U | 34%<br>**47%**<br>34% | **47%**<br>**47%**<br>**47%** | 34%<br>**47%**<br>66% | 0.51<br>0.29 | -3022.26 | 0.97 | 5 | -9.4 |

HSAR values (p estimates in a 3-by-3 grid) that share bold and/or italics font type were assumed to be equal for that model. P values are for rejection of the model in favor of the full model, by the likelihood ratio test with degrees of freedom equal to the difference in number of parameters (Par). Par = number of independent household transmission parameters simultaneously estimated. $\Delta$ AIC = difference in Akaike information criterion value versus the reference (Ref) model; a negative $\Delta$ AIC means the model is favored over the reference model, and the lowest $\Delta$ AIC model is favored over all the others.

**Table G.** Comparison of simpler age- and size-structured transmission models to most parsimonious model.

| | $\hat{p}_{ww2}$<br>$\hat{p}_{vw2}$<br>$\hat{p}_{uw2}$ | $\hat{p}_{wv2}$<br>$\hat{p}_{vv2}$<br>$\hat{p}_{uv2}$ | $\hat{p}_{wu2}$<br>$\hat{p}_{vu2}$<br>$\hat{p}_{uu2}$ | $\sigma_{34}$<br>$\sigma_{5+}$ | Log likelihood | P value | Par | $\Delta AIC$ |
| --- | --- | --- | --- | --- | --- | --- | --- | --- |
| Most parsimonious model | *34%*<br>**47%**<br>*34%* | **47%**<br>**47%**<br>**47%** | *34%*<br>**47%**<br>*66%* | 0.51<br>0.29 | -3022.26 | Ref | 5 | Ref |
| Equal HSAR except U-to-U | **44%**<br>**44%**<br>**44%** | **44%**<br>**44%**<br>**44%** | **44%**<br>**44%**<br>*66%* | 0.47<br>0.24 | -3024.70 | 0.03 | 4 | 2.9 |
| Equal HSAR for all ages | **54%**<br>**54%**<br>**54%** | **54%**<br>**54%**<br>**54%** | **54%**<br>**54%**<br>**54%** | 0.36<br>0.17 | -3029.35 | 0.0008 | 3 | 10.2 |
| No household size effect | *16%*<br>**30%**<br>*16%* | **30%**<br>**30%**<br>**30%** | *16%*<br>**30%**<br>*57%* | 1<br>1 | -3031.11 | 0.0001 | 3 | 13.7 |
| Equal HSAR for all ages and sizes | **27%**<br>**27%**<br>**27%** | **27%**<br>**27%**<br>**27%** | **27%**<br>**27%**<br>**27%** | 1<br>1 | -3053.50 | <0.0001 | 1 | 54.5 |

HSAR values (p estimates in a 3-by-3 grid) that share bold or italics font were assumed to be equal for that model. The “no household size effect” assumption fixes $\sigma_{34} = \sigma_{5+} = 1$ . P values are for rejection of the model in favor of the most parsimonious model, by the likelihood ratio test with degrees of freedom equal to the difference in number of parameters (Par). Par = number of independent household transmission parameters simultaneously estimated. $\Delta AIC$ = difference in Akaike information criterion value versus the reference (Ref) model; positive $\Delta AIC$ means the model is not favored over the reference model.

**Table H.**
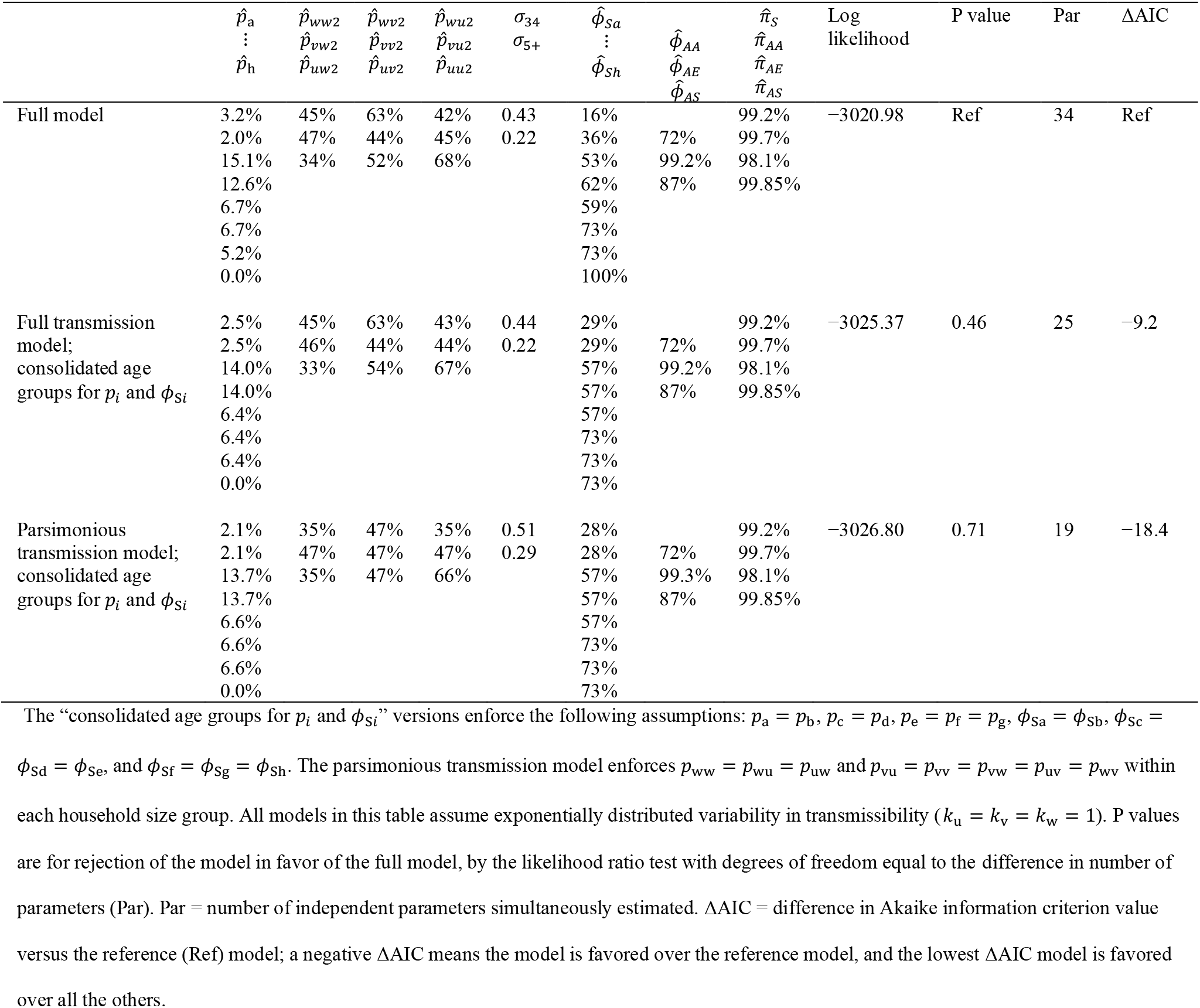
Consolidated age groupings for community acquisition and case ascertainment.

## References

1. Lloyd-Smith JO, Schreiber SJ, Kopp PE, Getz WM. Superspreading and the effect of individual variation on disease emergence. Nature 2005; 438(7066): 355–9.

2. VanderWaal KL, Ezenwa VO. Heterogeneity in pathogen transmission: mechanisms and methodology. Functional Ecology 2016; 30(10): 1606–22.

3. Pellis L, Cauchemez S, Ferguson NM, Fraser C. Systematic selection between age and household structure for models aimed at emerging epidemic predictions. Nat Commun 2020; 11(1): 906.

4. Buckee C, Noor A, Sattenspiel L. Thinking clearly about social aspects of infectious disease transmission. Nature 2021; 595(7866): 205–13.

5. Ma KC, Menkir TF, Kissler S, Grad YH, Lipsitch M. Modeling the impact of racial and ethnic disparities on COVID-19 epidemic dynamics. Elife 2021; 10.

6. Ferguson NM, Donnelly CA, Anderson RM. Transmission dynamics and epidemiology of dengue: insights from age-stratified sero-prevalence surveys. Philos Trans R Soc Lond B Biol Sci 1999; 354(1384): 757–68.

7. Meyerowitz EA, Richterman A, Gandhi RT, Sax PE. Transmission of SARS-CoV-2: A Review of Viral, Host, and Environmental Factors. Ann Intern Med 2021; 174(1): 69–79.

8. Qian H, Miao T, Liu L, Zheng X, Luo D, Li Y. Indoor transmission of SARS-CoV-2. Indoor Air 2020; ina.12766.

9. Yonker LM, Neilan AM, Bartsch Y, et al. Pediatric Severe Acute Respiratory Syndrome Coronavirus 2 (SARS-CoV-2): Clinical Presentation, Infectivity, and Immune Responses. J Pediatr 2020; 227: 45–52 e5.

10. Ghosh AK, Venkatraman S, Soroka O, et al. Association between overcrowded households, multigenerational households, and COVID-19: a cohort study. Public Health 2021; 198: 273–9.

11. Hart WS, Abbott S, Endo A, et al. Inference of the SARS-CoV-2 generation time using UK household data. Elife 2022; 11.

12. Endo A, Uchida M, Kucharski AJ, Funk S. Fine-scale family structure shapes influenza transmission risk in households: Insights from primary schools in Matsumoto city, 2014/15. PLoS Comput Biol 2019; 15(12): e1007589.

13. Cauchemez S, Carrat F, Viboud C, Valleron AJ, Boelle PY. A Bayesian MCMC approach to study transmission of influenza: application to household longitudinal data. Stat Med 2004; 23(22): 3469–87.

14. Bubar KM, Reinholt K, Kissler SM, et al. Model-informed COVID-19 vaccine prioritization strategies by age and serostatus. Science 2021; 371(6532): 916–21.

15. Metcalf CJ, Farrar J, Cutts FT, et al. Use of serological surveys to generate key insights into the changing global landscape of infectious disease. Lancet 2016; 388(10045): 728–30.

16. Longini I, Koopman JS. Household and community transmission parameters from final distributions of infections in households. Biometrics 1982; 38(1): 115–26.

17. Bi Q, Lessler J, Eckerle I, et al. Insights into household transmission of SARS-CoV-2 from a population-based serological survey. Nat Commun 2021; 12(1): 3643.

18. Toth DJA, Beams AB, Keegan LT, et al. High variability in transmission of SARS-CoV-2 within households and implications for control. PLoS One 2021; 16(11): e0259097.

19. Utah Health and Economic Recovery Outreach Home - Utah Hero. Available at: https://eccles.utah.edu/utah-hero/. Accessed 11/10/2023.

20. Samore MH, Looney A, Orleans B, et al. Probability-Based Estimates of Severe Acute Respiratory Syndrome Coronavirus 2 Seroprevalence and Detection Fraction, Utah, USA. Emerg Infect Dis 2021; 27(11): 2786–94.

21. Hershow RB, Wu K, Lewis NM, et al. Low SARS-CoV-2 Transmission in Elementary Schools - Salt Lake County, Utah, December 3, 2020-January 31, 2021. MMWR Morb Mortal Wkly Rep 2021; 70(12): 442–8.

22. Anti-SARS-CoV-2 ELISA (IgG) - Instructions for Use. Available at: https://www.fda.gov/media/137609/download. Accessed 10/20/2023.

23. SARS-CoV-2 IgG Architect - Instructions for Use. Available at: https://www.fda.gov/media/137383/download. Accessed 1/25/2021.

24. Dimension Vista SARS-CoV-2 IgG (COV2G) - Instructions for Use. Available at: https://www.fda.gov/media/145020/download. Accessed 10/20/2023.

25. Tang MS, Hock KG, Logsdon NM, et al. Clinical Performance of Two SARS-CoV-2 Serologic Assays. Clin Chem 2020; 66(8): 1055–62.

26. Rychert J, Couturier MR, Elgort M, et al. Evaluation of 3 SARS-CoV-2 IgG Antibody Assays and Correlation with Neutralizing Antibodies. J Appl Lab Med 2021; 6(3): 614–24.

27. Demmer RT, Baumgartner B, Wiggen TD, et al. Identification of Natural SARS-CoV-2 Infection in Seroprevalence Studies Among Vaccinated Populations. Mayo Clin Proc 2022; 97(4): 754–60.

28. Age Groups - MeSH - NCBI. Available at: https://www.ncbi.nlm.nih.gov/mesh/68009273.

29. Cronin CJ, Evans WN. Nursing home quality, COVID-19 deaths, and excess mortality. J Health Econ 2022; 82: 102592.

30. Paul LA, Daneman N, Schwartz KL, et al. Association of Age and Pediatric Household Transmission of SARS-CoV-2 Infection. JAMA Pediatr 2021; 175(11): 1151–8.

31. Bansal S, Grenfell BT, Meyers LA. When individual behaviour matters: homogeneous and network models in epidemiology. J R Soc Interface 2007; 4(16): 879–91.

32. Wegehaupt O, Endo A, Vassall A. Superspreading, overdispersion and their implications in the SARS-CoV-2 (COVID-19) pandemic: a systematic review and meta-analysis of the literature. BMC Public Health 2023; 23(1): 1003.

33. Ke R, Martinez PP, Smith RL, et al. Daily longitudinal sampling of SARS-CoV-2 infection reveals substantial heterogeneity in infectiousness. Nat Microbiol 2022; 7(5): 640–52.

34. Davies NG, Klepac P, Liu Y, et al. Age-dependent effects in the transmission and control of COVID-19 epidemics. Nat Med 2020; 26(8): 1205–11.

35. Keegan LT, Truelove S, Lessler J. Analysis of Vaccine Effectiveness Against COVID-19 and the Emergence of Delta and Other Variants of Concern in Utah. JAMA Netw Open 2021; 4(12): e2140906.

36. Madewell ZJ, Yang Y, Longini IM, Jr., Halloran ME, Dean NE. Household Secondary Attack Rates of SARS-CoV-2 by Variant and Vaccination Status: An Updated Systematic Review and Meta-analysis. JAMA Netw Open 2022; 5(4): e229317.

